# Temporary Shock or Lasting Scar? Life Expectancy Trajectories Since COVID-19

**DOI:** 10.64898/2026.02.25.26347112

**Authors:** Jennifer B. Dowd, Jonas Schöley, Antonino Polizzi, José Manuel Aburto, Hannaliis Jaadla, Haohao Lei, Ridhi Kashyap

## Abstract

The COVID-19 pandemic led to substantial global life expectancy losses. Historically, life expectancy reversals were followed by rapid returns to previous trajectories, but it remains unclear whether this pattern applies to COVID-19. We extend life expectancy estimates through 2024 for 34 high-income countries and quantify annual and cumulative life expectancy deficits by comparing observed life expectancy to counterfactual forecasts based on pre-pandemic trajectories. Five years after the pandemic’s onset, recovery remains incomplete: in 2024, 31 of 34 countries still had lower-than-expected life expectancy. We identify four trajectories: (a) early deficit (largest deficits in 2020 or 2021 with gradual recovery); (b) second wave peak (largest deficits in 2021 with a sharper rebound); (c) late deficit (minimal early impact followed by smaller deficits from 2022 onward); (d) prolonged deficit (smaller but persistent deficits without a sharp peak). In general, countries with severe second-wave peaks (such as the USA and Bulgaria) had the largest 5-year deficits. In contrast, countries that delayed widespread infection (e.g., Norway, Japan) saw later deficits that persisted through 2024, but with lower overall mortality. Our findings suggest that COVID-19 was not a uniform, short-lived mortality shock. Instead, most high-income countries experienced multi-year disruptions, with varied recovery patterns that continue to shape population health five years on.

## Introduction

The COVID-19 pandemic produced the largest declines in life expectancy in many high-income countries since World War II. In 2020 and 2021 alone, life expectancy declined by more than one year in many countries, reversing decades of steady improvement (Schöley et al. 2022). Historically, life expectancy reversals have been short-lived, with life expectancy returning quickly to its prior trajectory, an assumption already made by the most recent 2024 revision of the United Nations World Population Prospects (United Nations, Department of Economic and Social Affairs, Population Division 2024). Whether the COVID-19 pandemic will follow a similar pattern of a temporary mortality shock or a more lasting disruption to life expectancy trends is not known.

Recent work examining life expectancy reversals since 1950 suggests that mortality shocks do not typically alter long-run trajectories (Goldstein and Lee 2024). Yet most prior temporary declines in life expectancy occurred during decades of rapid gains, which could obscure the negative impact of smaller shocks on life expectancy trajectories. Recently, these steady patterns of life expectancy improvements have slowed in many high-income countries (Dowd et al. 2024). Thus, the emergence of SARS-CoV-2 as a novel pathogen comes at a time when improvements in life expectancy are becoming more difficult to attain, which may increase the salience of mortality shocks.

For any mortality shock, competing mechanisms can imply different long-term outcomes. Goldstein and Lee (2024) find that declines in life expectancy are often followed by faster-than-expected increases, especially at older ages. This may be due to mortality displacement or selective mortality of the frailest (Andersson and Lindholm 2022; Chen et al. 2026), leaving survivors with better-than-average health. Alternatively, biological “scarring,” ongoing infection risk, health system disruption, and broader social changes could worsen the health of survivors in the future. While selective mortality of the most frail is plausible due to the steep age profile of COVID-19 mortality (Dowd et al. 2020), COVID-19 infection may also increase cardiovascular disease and mortality risk among survivors (Xie et al. 2022; Bruno et al. 2025). Thus, the balance between scarring and selection, and their impact on life expectancy recovery, is not obvious a priori. What’s more, SARS-CoV-2 is a novel human pathogen, adding to the toll of common respiratory viruses that already pose an ongoing mortality risk. Besides the direct impacts of COVID-19 infection on mortality, the pandemic created dramatic social disruptions that likely altered patterns of mortality from other causes in ways that differ across countries (Polizzi et al. 2024). Given the magnitude and complexity of COVID-19 as a mortality shock, it is unclear whether COVID-19 will follow the expected pattern of rapid “bounce backs” to pre-pandemic mortality trajectories or whether it reflects an additional and ongoing mortality risk that alters those trajectories. This distinction has significant implications beyond demographic accounting. If COVID-19 is a transient shock, the long-run upward trajectories of population health and longevity remain intact. If the pandemic has more lasting impact, countries could face policy challenges related to a persistently higher burden of chronic disease and premature mortality to which health and economic systems must adapt.

We test this question by updating life expectancy through 2024 for 34 high-income countries and comparing observed values to counterfactual forecasts based on pre-pandemic trends We call this difference between counterfactual and observed levels “life expectancy deficits.” By quantifying annual and cumulative life expectancy deficits, we evaluate whether countries have returned to their expected trajectories five years after the onset of the pandemic. We find that most countries have not. Rather than a uniform rebound, we find distinct patterns in the timing, magnitude, and persistence of life expectancy deficits across countries.

### Data and Methods

We calculate life tables and associated life expectancies by sex from 2000 through 2024 for 34 high-income countries (Austria, Belgium, Bulgaria, Chile, Croatia, Czech Republic, Denmark, England & Wales, Estonia, Finland, France, Germany, Greece, Hungary, Iceland, Israel, Italy, Japan, Latvia, Lithuania, Luxembourg, Netherlands, Northern Ireland, Norway, Poland, Portugal, Scotland, Slovakia, Slovenia, Spain, Sweden, Switzerland, Taiwan, and the USA). For the individual years 2020 through 2024, we calculate counterfactual life expectancies based on the continuation of pre-pandemic mortality trends and derive associated life expectancy deficits – the difference between observed and counterfactual life expectancy – as an age-standardized and trend-adjusted measure of excess mortality. In addition, we calculate the overall 5-year life expectancy deficit by pooling deaths and person-years of exposure across 2020-2024 and constructing an aggregated 5-year life table.

All estimates are based on midyear population and death counts by age, sex, and country sourced from the Short Term Mortality Fluctuations Database (HMD-STMF) (Jdanov et al. 2021), the Human Mortality Database (HMD) (Barbieri et al. 2015; Max Planck Institute for Demographic Research et al. 2025), the UN World Population Prospects (WPP-2024) (United Nations Department for Economic and Social Affairs 2025), and national statistical offices (ONS, CDC). For a complete list of data sources by country, see the methods appendix “Data sources.” The term “country” throughout the paper refers to the populations for which life expectancy is estimated and does not imply a statement about the legal status of any territory.

We selected countries represented in the Human Mortality Database for which there were reliable death counts by age and sex for 2024 from either the HMD’s annual data series or HMD-STMF as of February 5^th^ 2026, the date on which we froze our source data. Included countries are a) designated as “high-income” by the World Bank’s income classification, and b) have a highly reliable data series, as determined by the ongoing documentation, validation, and harmonization work of the Human Mortality Database.

Age-specific death counts are harmonized to a single year age grouping with open age group 100+ using the Penalized Composite Link Model (PCLM) method (Rizzi et al. 2015; Pascariu et al. 2018). Weekly death counts are aggregated to annual counts, and midyear population estimates are converted to person-years of exposure, adjusting for the effect of leap weeks. To adjust for systematic bias in registered deaths between annualized STMF data and HMD annual totals, we estimate correction factors based on a statistical model (see methods appendix “Bias correction of age-specific annual death counts”). Throughout the analysis, population exposures and annual death counts by sex are aggregated as needed to derive results for both sexes combined.

We calculate period life tables using the standard piecewise-exponential model with harmonized annual death counts and person-years at risk of death (hereafter “population exposures”) by age and sex. We then fit a Poisson-Lee-Carter model (Lee and Carter 1992) using the R library StMoMo (Villegas et al. 2015) to the available pre-pandemic data from 2000 to 2019 and forecast counterfactual age-specific mortality rates and death counts from 2020 through 2024. We decompose the life expectancy deficits into age-specific contributions using the Arriaga method (Arriaga 1984). Life expectancy deficits are then derived along with excess mortality P-scores (Ullrich-Kniffka and Schöley 2024) and the Mean unfulfilled lifespan (MUL) as alternative measures of excess mortality (Heuveline 2023). These alternative measures were highly correlated with our estimate of life expectancy deficits (see methods appendix), and we focus on life expectancy deficits in the main manuscript. The clustering of countries into typologies of life expectancy trajectories was estimated with a hierarchical clustering algorithm implemented in the R package dtwlust (Alexis Sarda-Espinosa 2024) (see methods appendix “clustering of life expectancy deficits” for details).

The Lee-Carter method used to estimate counterfactual life expectancy rests on several key assumptions. It is extrapolative, projecting mortality trends based on the training period. Rates of mortality improvement are allowed to vary by age, but the age pattern of improvements is assumed to be fixed over time (Basellini et al. 2023). We conducted sensitivity analyses using a smaller window (2010-2019) of baseline data for our Lee-Carter forecasts, as well as two alternative forecasting approaches with different assumptions (see methods appendix “robustness and sensitivity”).

We perform simulation-based uncertainty propagation throughout our analysis via a Poisson parametric bootstrap of the life table estimates and a random walk simulation of the Lee-Carter forecast. Based on this, we derive a p-value for the estimated life expectancy deficit, i.e. the probability of observing or exceeding an estimated life expectancy deficit given the continuation of pre-pandemic trends (see methods appendix, “Life table inference” for more detail). The complete analysis workflow is explained in detail in the methods appendix, and reproducible data and code for all analyses are available at (Jonas Schöley 2026): https://doi.org/10.5281/zenodo.21808782.

## Results

Figure 1 shows annual life expectancy estimates from 2010 through 2024 (dots) compared with Lee-Carter estimates for 2020 through 2024, assuming continued pre-pandemic mortality trends (line with 95% prediction intervals). Corresponding numeric estimates for each year with 95% prediction intervals are shown in Table S1. As of 2024, the mean LE deficit was negative in 31 out of 34 countries, indicating that life expectancy levels had not returned to their pre-pandemic trajectories five years later. The negative deficits were statistically significant at the 5% level in 12 countries in 2024: Austria (−0.68 years, p=0.020), Chile (−0.51, p=0.020), Denmark (−0.40, p=0.048), Finland (−0.64, p<0.001), Greece (−0.78, p=0.016), Israel (−1.23, p<0.001), Japan (−1.07, p<0.001), Netherlands (−1.08, p<0.001), Northern Ireland (−1.33, p<0.001), Norway (−0.78, p<0.001), Taiwan (−1.04, p<0.001), and USA (−0.55, p<0.001). Across the overall 2020-24 period, the five-year deficit was significantly negative in all analysed countries, except Iceland and Luxembourg (Table S1).

**Figure 1.**
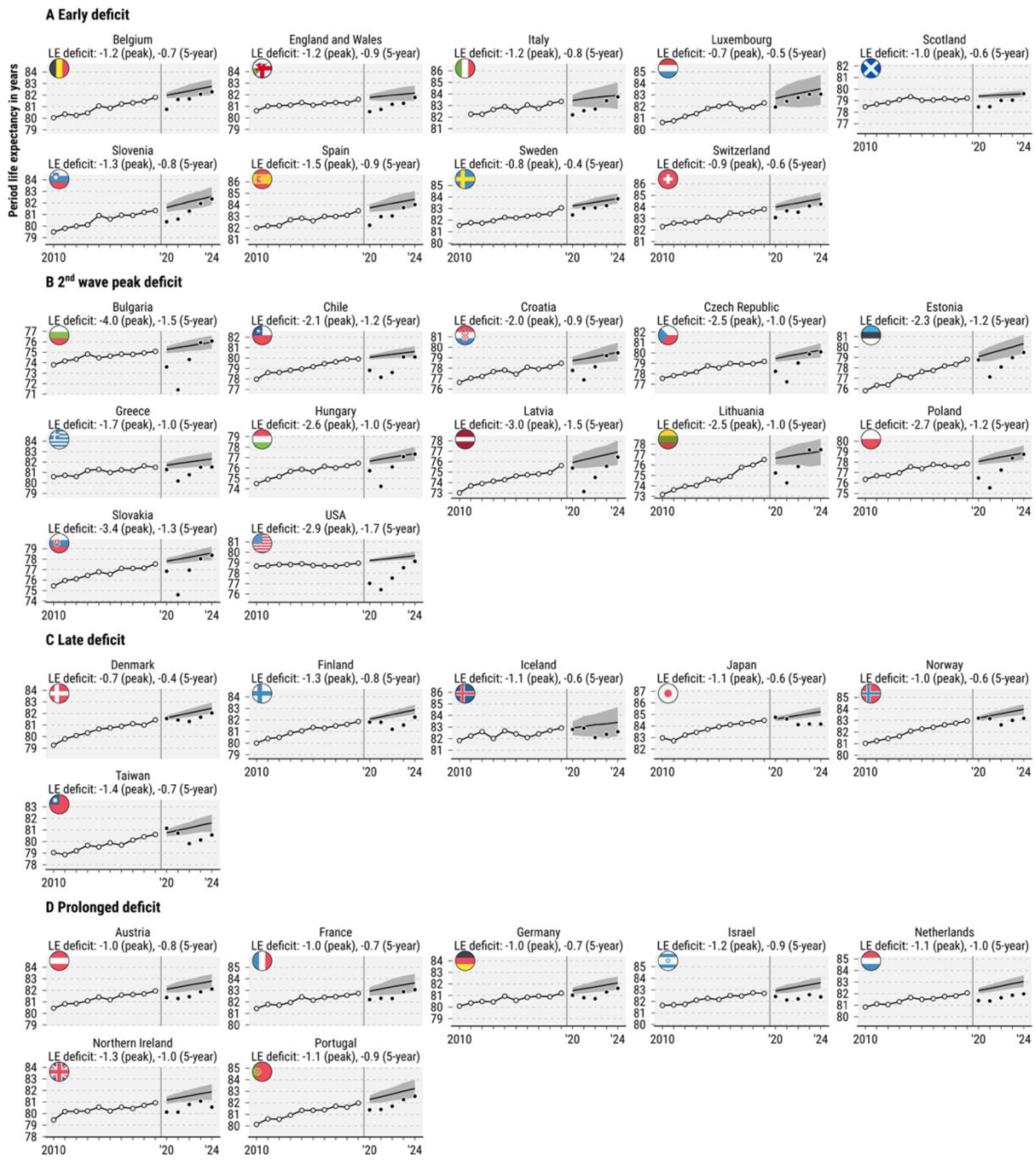
Period life expectancy at birth trends 2010-24. With the start of the pandemic in 2020, we superimpose observed life expectancy (black dots) over the life expectancy forecast based on pre-pandemic trends (black line surrounded by 95% prediction intervals). Countries are grouped into four typologies of mortality trajectories.

We identify four broad typologies of mortality experiences based on our clustering analysis, shown for individual countries in Figure 1 and grouped in Figure 2. Countries that experienced their worst mortality peaks in 2020 or 2021, without pronounced worsening in 2021 (“early deficit”), include Belgium, England and Wales, Italy, Luxembourg, Scotland, Slovenia, Spain, Sweden, and Switzerland. This group saw a peak deficit of −1.09 years of life expectancy on average, and an average −0.68-year deficit across the entire period. Countries that experienced their sharpest mortality losses in 2021 (“second wave peak”) include most countries of Eastern Europe plus Chile, Greece, and the USA. This group had both the highest average peak deficit (−2.65 years) and overall five-year deficit (−1.20 years) of the four typologies. The third group (“late deficit”) includes countries that delayed substantial mortality losses until 2022, with substantial but not full recovery by 2024 (Iceland, Denmark, Finland, Japan, Norway and Taiwan). This group had an average peak deficit of −1.09 years and an overall deficit of −0.62 years. Finally, seven countries showed no definitive pandemic “peak” but persistent life expectancy deficits across the whole period (Austria, France, Germany, Israel, the Netherlands, Northern Ireland, and Portugal) (“prolonged deficit”).

**Figure 2.**
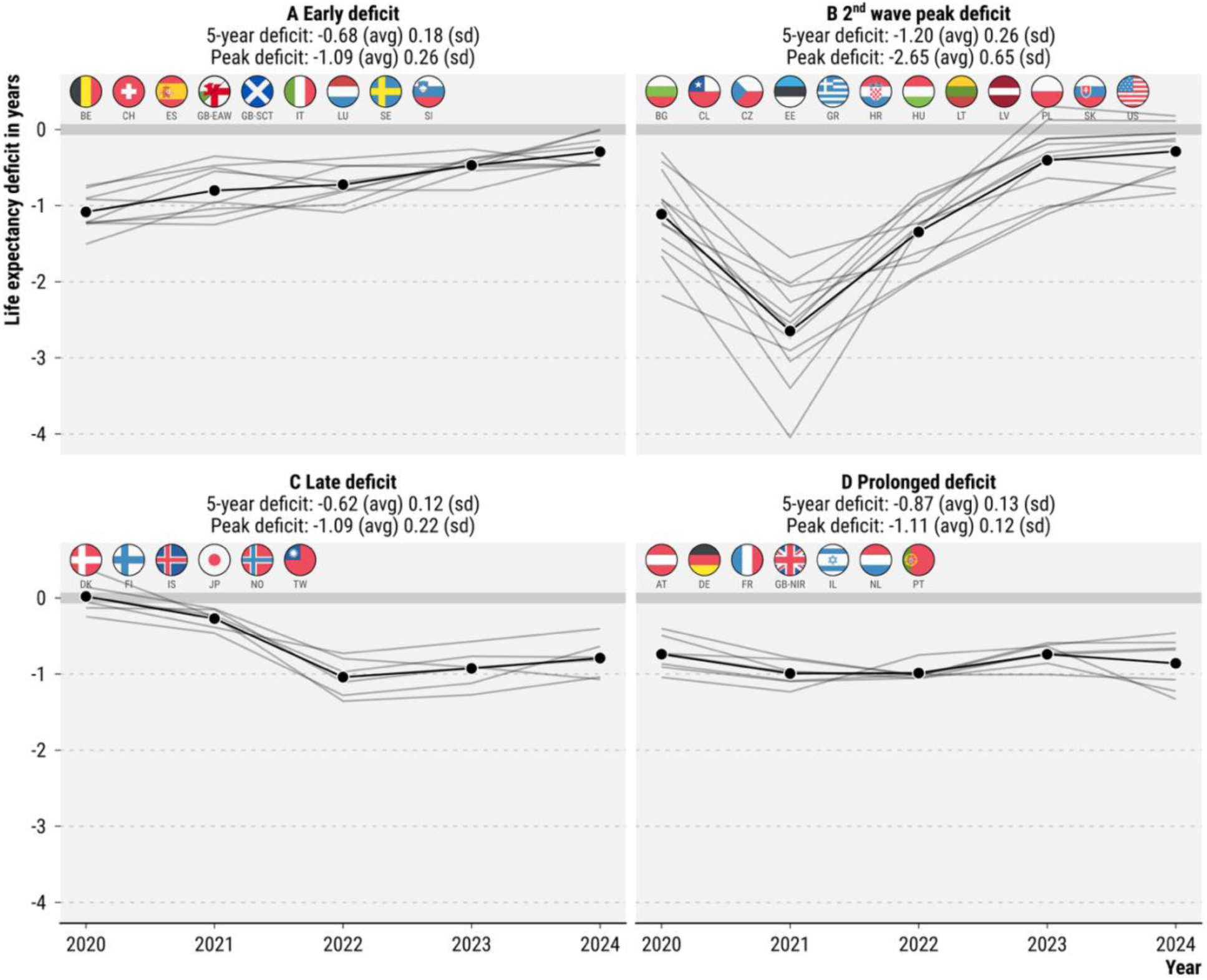
A typology of period shocks along life expectancy deficit patterns 2020 through 2024. Each grey line represents the annual life expectancy deficit in years (actual minus expected life expectancy) in each of the 34 countries. The black line depicts the average deficit for that group of countries. Annotated for each panel are the mean (“avg”) and standard deviation (“sd”) across countries of the deficit at peak and the five-year deficit.

The “prolonged deficit” group had the second-highest average peak deficit (−1.11 years) and overall 5-year deficit (−0.87 years). While these typologies do not fit each individual country perfectly, they effectively summarize the broad patterns and major variations in the timing, severity, and recovery of life expectancy. Figure 2 also demonstrates that life expectancy deficits persisted for multiple years in most countries rather than rebounding immediately after their peak shock.

Figure 3 shows country rankings of life expectancy deficits for each individual year and the overall 2020-24 period. While countries that saw the most severe peaks have largely bounced back near projected life expectancy levels in 2024, their total life expectancy deficits across the overall period were among the largest, for example a −1.49 (p<0.001) year deficit over the five-year period for Bulgaria, and a −1.73 (p<0.001) year deficit for the USA. Countries that delayed significant COVID-19 mortality until 2022 generally had lower cumulative deficits, such as Denmark (−0.43 years, p<0.001), Norway (−0.56, p<0.001), and Japan (−0.57 years, p<0.001) as did countries with early peaks but minimal losses thereafter, e.g. Sweden (−0.41, p<0.001) or Switzerland (−0.63, p<0.001).

**Figure 3.**
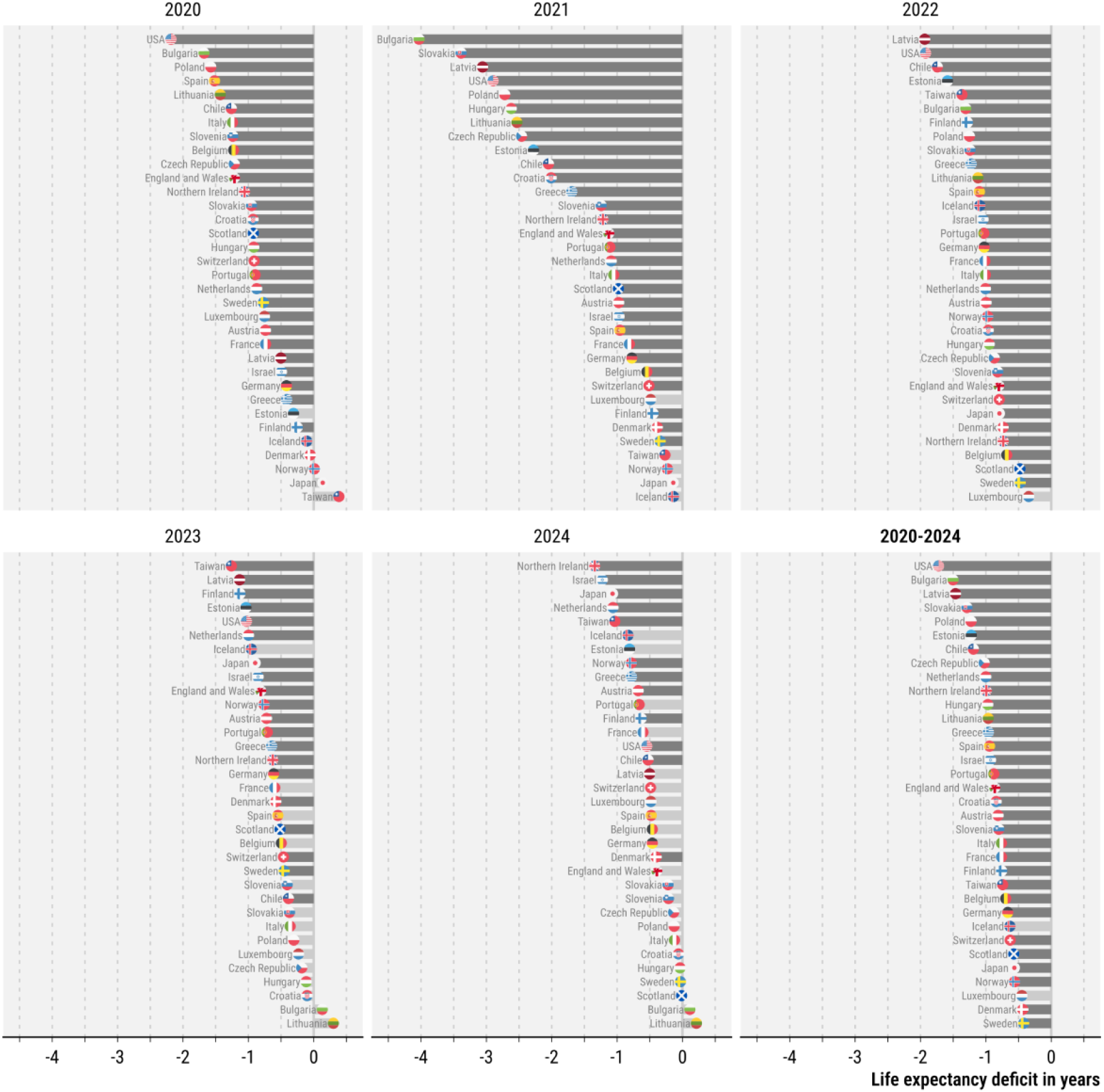
Country rankings of life expectancy deficits, 2020-2024. Shown are the differences in life expectancy at birth between the actual and expected period life tables across 34 countries. Dark grey bars indicate a significant life expectancy deficit (p≤0.05), while light grey bars indicate a non-significant deficit or no deficit. The lower-right panel shows life expectancy deficits for the total five-year period 2020 through 2024.

Some countries, such as the Netherlands (−1.01 years, p<0.001), Germany (−0.66, p=0.004), and Portugal (−0.89, p<0.001), experienced less sharp but persistent deficits that accumulated to significant losses over the full 5-year period. As a country with one of the earliest COVID-19 waves and a sizeable mortality shock in 2020 (−1.24 years, p<0.001), Italy saw substantial recovery in later years leading to a less dramatic overall 5-year deficit (−0.77 years, p=0.02), similar to the overall deficit of England and Wales (−0.87, p<0.001). Among these 34 high-income countries, the USA had the largest overall life expectancy deficit for the 2020-24 period (−1.73, p<0.001), followed by Bulgaria (−1.49, p<0.001) and Latvia (−1.47, p<0.001). The USA recorded life expectancy deficits in each individual year and had the largest deficit among all countries in 2020. The USA life expectancy deficit ranked among the largest through 2023, with its 2024 shortfall improving towards the middle of the pack.

**Figure 4** shows the age-specific decomposition of annual life expectancy deficits for each country. If mortality displacement were an important driver of rapid rebounds, this might appear as a “mirroring” of age patterns, with high mortality in early periods showing faster recovery in later periods. We observe evidence of such mirroring in Bulgaria, Croatia, Hungary, Lithuania, and Slovakia, with higher-than-expected life expectancy gains at ages 80+ since 2023. In Iceland, Japan, and Taiwan, we see evidence of “reverse” displacement, with higher-than-expected life expectancy at the oldest ages in the first year of the pandemic. This is likely due to minimal COVID-19 spread in these countries early in the pandemic, combined with extra precautions taken to protect older people, which reduced typical mortality risks from influenza, pneumonia, and accidents (Groves et al. 2021).

**Figure 4.**
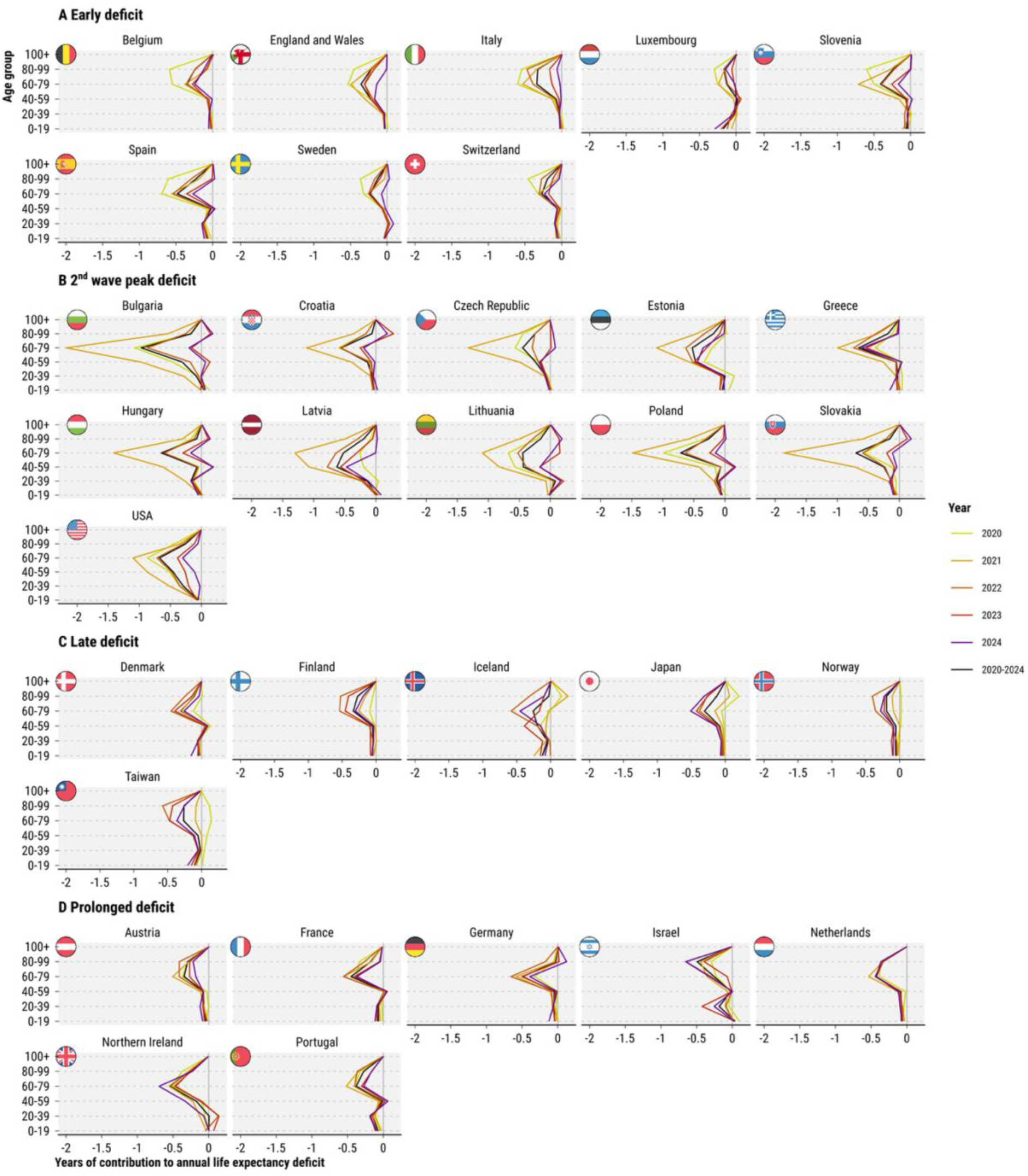
Age-specific contributions to annual life expectancy deficits 2020 through 2024 and for all years combined. Note Scotland and Chile are excluded from age decomposition due to data issues.

The four typologies of pandemic mortality are also reflected in the pattern of age-specific life expectancy deficits. Most notably, many of the “second wave peak” countries with the largest life expectancy deficits saw significant losses in younger age groups (40-59 years) compared to “early deficit” countries, where deficits were more concentrated above age 60. This pattern of mid-life mortality was especially prominent in the USA and Eastern Europe (Bulgaria, Czech Republic, Estonia, Poland). Patterns of recovery in these younger age groups did not show a mirroring effect, although there were some signs of a reduction in deficits over time. This pattern suggests that factors other than mortality displacement were shaping total deficits and life expectancy trajectories. Persistent LE declines in younger age groups after 2021 contributed to overall declines in life expectancy, even as older-age mortality recovered more quickly.

Results by sex are available in the supplementary Figures S1-5. In general, life expectancy deficits were larger for males compared to females, with the USA and Latvia showing particularly large sex differences. Age contributions to life expectancy deficits were largely symmetric between the sexes with the notable exception of Israel where excess mortality among males of drafting age stands out in 2023 and 2024. Our estimates are consistent with evidence that COVID amplified pre-existing sex differences in all-cause mortality, but the magnitude varied across countries, highlighting the role of social context in shaping sex-specific pandemic vulnerability (Doniec et al. 2026).

### Robustness Checks

We checked the sensitivity of our estimates to different counterfactual mortality forecasting assumptions, including a 10-year rather than 20-year fitting window for our Lee-Carter models. Our central conclusions regarding the typology of pandemic trajectories, general rank ordering of countries, and persistence of estimated deficits in 2024 were consistent across different modeling choices. The USA was more sensitive to the choice of fitting period, due to its pronounced life expectancy stagnation since 2010. A 10-year fitting window continues an almost flat US trajectory, leading to lower life expectancy deficits, whereas a 20-year fitting window extrapolates a longer-term trend of slight life expectancy increase. While the US had the largest 5-year deficit using the 20-year fitting period, it ranked 7^th^ worst using the 10-year fitting period (after Latvia, Bulgaria, Lithuania, Slovakia, Chile, and Estonia). Despite this relative improvement for the USA, it’s notable that the overall magnitude of USA deficits did not change dramatically, and the US remained among the countries with the highest deficits in 2020, 2021, and 2022 despite having the lowest expectations for counterfactual mortality improvements. Our results were also consistent across different forecasting assumptions, including life expectancy extrapolation and linear extrapolations of log age-specific death rates. Full details can be found in the “robustness and sensitivity” section of the methods appendix.

## Discussion

Five years on, the COVID-19 pandemic does not look like a short-lived mortality shock. Instead, it represents a multi-year disruption to life expectancy trajectories in most high-income countries. In 31 of 34 countries we analyzed, life expectancy in 2024 remained below levels expected based on pre-pandemic forecasts. Recent global population projections, including the United Nations World Population Prospects 2024 revision, assumed that life expectancy has returned to its pre-pandemic trajectory (United Nations, Department of Economic and Social Affairs, Population Division 2024). Our findings suggest that this assumption may be premature for many high-income countries.

In general, countries that experienced the most severe life expectancy deficits in the first two years of the pandemic were more likely to have returned to their pre-pandemic trajectories by 2024, but with larger 5-year life expectancy deficits. While this pattern most resembles the transient shock and quick recovery pattern described by Goldstein and Lee (2024), we observe a wider range of mortality experiences across our 34 countries. Almost all countries experienced life expectancy deficits that persisted for multiple years, suggesting that COVID-19 was a more significant mortality shock than the typical post-1950 life expectancy reversals described by Goldstein and Lee. Previous analysis of life expectancy declines since 1900, including the 1918 flu, found that life expectancy levels generally recovered to pre-crisis levels in one or two years (Schöley et al. 2022), though Silva and Aburto found evidence of longer recovery times from major historical European cholera pandemics (Silva and Aburto 2025).

A central question is not simply whether life expectancy has rebounded to pre-pandemic levels, but whether the pandemic has altered longer-run trajectories of mortality improvement. Figure S6 illustrates three stylized scenarios. In scenario A, the pandemic induces a temporary shock followed by full recovery in both level and slope of life expectancy, as described by Goldstein and Lee. In scenario B, countries return to their pre-pandemic rate of improvement but at a lower level due to persistent additive mortality risk from COVID-19. In scenario C, the pandemic induces a structural break in the slope of mortality decline. In this most pessimistic scenario, the pandemic marks a fundamental shift in the drivers of mortality that slow improvements in the longer run. This could be through direct biological mechanisms, such as post-acute sequelae of COVID (Cai et al. 2024), or “Long COVID,” that increase vulnerability to other chronic diseases. Incomplete recovery could reflect ongoing pressure on health systems that were strained during COVID, disruptions to medical and research innovation, and increased political polarization (Nuzzo 2026). If COVID-19 slows the rate of mortality improvement in the longer run, this has much bigger downstream consequences for economies and health systems than a shorter period of “lost” progress. In the best-case scenario, both the slope and level of life expectancy will fully return to pre-pandemic trajectories, as patterns in several Eastern European countries already imply. In this scenario, the lives lost prematurely during the COVID-19 pandemic are still tragic, but there is no lost progress or ongoing elevated mortality risk for survivors. While it is too early to distinguish conclusively between these scenarios, the persistence of deficits in many countries raises the possibility that COVID-19 may represent either a more persistent level shift or a slowdown in mortality improvement.

The typology of pandemic mortality trajectories we identify provides insight into how countries may be sorting into these different long-term scenarios. Among the “second wave peak” group, only the Eastern European countries saw a full recovery to pre-pandemic life expectancy, while the USA (−0.55 years, p<0.001), Chile (−0.51 years, p=0.02), and Greece (−0.78 years, p=.016) had still not returned to expected 2024 levels. This heterogeneity in bounce-back among countries with the sharpest losses may reflect differences in their longer-term mortality dynamics. In most Eastern European countries, life expectancy began from lower levels in 2010 and improved faster than in other high-income countries, reflecting recovery from the post-Soviet mortality crisis and delayed benefits from the “cardiovascular revolution” (Aburto and van Raalte 2018; Fihel and Pechholdová 2017). On the one hand, a steeper pre-pandemic trajectory means that returning to expected levels in 2024 is a greater reach than for countries whose improvements were stagnating prior to the pandemic. On the other hand, the steep upward life expectancy trajectories in Eastern Europe prior to COVID-19 may reflect favourable underlying trends and the potential for unrealized mortality gains that can more easily buffer an acute mortality shock. The USA, by contrast, started at higher life expectancy levels than the Eastern European countries in 2010, but saw virtually no improvements in the decade prior to the pandemic. While this means the USA has a lower bar to return to its pre-pandemic trajectory, it also lacks the upward momentum of favourable mortality trends that could compensate for the COVID-19 shock. Mortality displacement is another potential explanation for faster recovery after the largest mortality shocks. There is some evidence of displacement (better-than-expected gains in years after losses) at older ages for Bulgaria, Croatia, Hungary, Lithuania, and Slovakia. Bulgaria, Hungary, and Poland, also saw faster-than-expected gains in 2023 and 2024 for people ages 40-59. Mortality displacement is less likely in this age group, suggesting perhaps a return to rapid pre-pandemic improvements that are outperforming the Lee-Carter forecasts.

Countries in the “early deficit” group were also likely to have rebounded close to their pre-pandemic life expectancy trajectories by 2024, though the bounce-back was not as definitive as in Eastern Europe. Italy, for example, had fully recovered by 2024 (−0.14, p=0.396) while Spain (−0.46, p=0.104) and England & Wales (−0.39, p=0.096) had lasting but marginally significant deficits. This less pronounced recovery may reflect the specific nature of “early peak” country experiences. Despite the high mortality of initial waves in the Spring of 2020, mortality was generally more localized (i.e. Lombardy, Italy) without affecting the country as a whole. In response to severe first waves, countries often implemented stringent COVID-19 policy measures that drove transmission to low levels and mitigated further life expectancy losses in 2021. When COVID-19 restrictions were largely lifted after the vaccine rollout in 2021, infection-naive populations experienced some excess mortality as widespread COVID-19 infections hit the population for the first time. This “early deficit” experience thus closely resembles the lower overall 5-year mortality burden of “delayed deficit” countries (such as Japan, Norway, Taiwan), who avoided widespread COVID-19 transmission until 2022. While these “delayed deficit” countries could not avoid COVID-19-associated mortality indefinitely, their overall mortality burden was significantly lower than the worst-case “second wave peak” countries, which experienced widespread transmission much earlier.

One mechanism that could drive quick life expectancy bounce-backs, like scenario A, is mortality displacement. If COVID-19 deaths primarily pulled forward the timing of deaths among frail individuals, we would expect lower-than-expected mortality in subsequent years. We find limited evidence for such mortality displacement, primarily at older ages in a subset of Eastern European countries. However, ongoing COVID-19 circulation and indirect effects of the pandemic could offset any compositional advantages among survivors, making displacement difficult to detect in aggregate data. For example, Sweden’s first-wave mortality was highly concentrated among older adults, especially nursing home residents. Using linked individual-level administrative data from Sweden, Ebeling, et al. estimated that more than half of all COVID-19 deaths in 2020 had a remaining life expectancy of less than 4 years, a time frame within which we might then expect to see evidence of displacement (Ebeling et al. 2022). Despite this finding, our age-specific results for Sweden don’t show strong evidence for mortality displacement at older ages, perhaps due to the compensating additional risk among survivors.

While most countries experienced reductions in excess mortality at older ages over time, likely due to protection from vaccination and other precautions, our age decompositions revealed persistent life expectancy deficits at older ages relative to pre-pandemic trends (apart from Bulgaria, Croatia, Hungary, Lithuania, and Slovakia). This suggests that, in general, COVID-19 was an additive rather than a competing risk for mortality over this period. Formal demographic modelling suggests that the impact of COVID-19-induced mortality selection on population death rates is likely to be minimal, partly because even large mortality shocks don’t kill a large enough fraction of the population to change the composition, and individual deaths tend to be unpredictable (Goldstein, under review, (Einav et al. 2018). This is consistent with recent excess mortality estimates that found evidence for COVID-19 pandemic mortality displacement in only 3 of 34 countries (Greece, Latvia, and Poland) at ages 85 and over (Chen et al. 2026), and earlier work that estimated an average of 10-11 years of life lost per COVID death (Hanlon et al. 2021; Goldstein and Lee 2020)

While we estimate life expectancy deficits as a measure of overall COVID-19 mortality burden, it is difficult to make strong causal claims about the specific policies or other factors that led to mortality differences across countries. In a previous paper, we found that higher COVID-19 vaccination uptake by October 2021 was strongly correlated with smaller life expectancy deficits in the last quarter of 2021 (Schöley et al. 2022). Further evidence showed that European countries with higher and faster COVID-19 vaccination rates had lower excess mortality through May 2022, but these countries also had lower excess mortality *prior* to the vaccine rollout, suggesting that vaccination rates are correlated with other key country-level characteristics (Matveeva and Shabalina 2023). Pre-existing country differences in population health and in risk factors such as obesity and diabetes may have contributed to greater population-level vulnerability to COVID-19 mortality in countries like the USA, which entered the pandemic with already concerning levels of midlife mortality (Martinson et al. 2022; Bishop et al. 2022; Dowd et al. 2023).

Our results do not necessarily tell one coherent story about the success or failure of COVID-19 mitigation measures and mortality at the country level. The poor country ranking of most Eastern European countries and the USA is consistent with less stringent COVID-19 policies in 2020 and 2021 and lower vaccination uptake compared to most Western European countries (Matveeva and Shabalina 2023; Smyth et al. 2022). But other country-specific paths are less expected. Sweden for example, suffered the largest losses of the Nordic countries in 2020, but had among the lowest overall deficits across the entire 2020-2024 period. Sweden received significant attention early in the pandemic for its more relaxed approach to COVID-19 mitigation compared to other European countries (Vogel 2021). While Sweden’s low overall pandemic mortality is notable, it is unlikely that we can infer much about the relative effectiveness of COVID-19 mitigation strategies from this strong performance. In practice, Sweden’s COVID-19 response differed only in the earliest weeks of the pandemic, with later policies more similar to those of other Nordic countries (Lindström 2023; Hallberg et al. 2025). Overall, identifying the key policy and other drivers of differences in COVID-19 mortality across countries is a challenging but important ongoing research agenda (Nuzzo and Ledesma 2023).

Whether countries ultimately return to pre-pandemic trajectories will depend on the mechanisms that shape post-pandemic mortality. Longer-term cohort scarring could produce a sustained level shift or a slowdown in mortality improvement. Previous pandemic pathogens have had long-term impacts on the health of affected cohorts. People exposed in utero to the 1918 influenza pandemic had elevated mortality at older ages (Myrskylä et al. 2013). Significant effects of prenatal COVID-19 infections on infant health have already been found (Torche and Nobles 2023), and some evidence suggests that COVID-19 infections may raise the risk of subsequent cardiovascular disease (Xie et al. 2022). Given these potential lasting risks, population scientists have called for careful monitoring of the pandemic’s impacts on life-course health and aging for decades to come (Easterlin et al. 2021). If such effects persist at scale, they could slow the rate of mortality improvement, shifting countries toward a structural change in life expectancy trajectories rather than a temporary deviation.

The age distribution of life expectancy deficits reinforces concerns about indirect effects of the pandemic. In many “second wave peak” countries, including the USA and Eastern Europe, excess mortality at ages 40-79 contributed substantially to large overall deficits. Although our analysis cannot distinguish causes of death, the excess mortality in these age groups suggests a potential role for non-COVID-19 causes, as identified by Polizzi et al. (2024). Mortality attributed to cardiovascular disease (CVD) worsened in many countries during the pandemic, including those with previously steadily improving CVD mortality (e.g. Eastern Europe), as well as those with stagnating improvements in CVD (e.g. USA), though some of this likely reflects misattributed COVID-19 deaths (Paglino et al. 2024; Polizzi et al. 2024). Increases in drug-related mortality were particularly notable in the USA, while increases in alcohol-related mortality were seen in many Eastern European countries (Rehm et al. 2024). Israel also experienced significant life expectancy deficits at younger age, especially for men, likely reflecting war-related mortality emerging in 2023, alongside older age life expectancy deficits more consistent with COVID-19^1^. Continued monitoring of age and cause-specific trends can help assess whether these effects were transitory or contribute to longer-term shifts in mortality trajectories.

### Strengths and limitations

This study provides the first comprehensive estimate of changes in life expectancy through 2024 to assess whether high-income countries have returned to their pre-pandemic trajectories. While period life expectancy is a standard way to track and compare mortality conditions across countries and time for a hypothetical cohort, it can be challenging to interpret when mortality conditions are changing (Luy et al. 2020). Heuveline suggested that during mortality shocks, period life expectancy deficits can be interpreted as the age-standardized value of the average lifespan reduction for people dying during the mortality shock (Heuveline 2023). We compared our LE deficits to Heuveline’s suggested “Mean unfulfilled lifespan” measure – an average of remaining life expectancy weighted by age-specific excess deaths – and found the metrics to be highly correlated, although our LE deficits tended to be a few months lower on average (see methods appendix, “Alternative measures of excess mortality”). We apply a similar interpretation to our estimated life expectancy deficits and argue that life expectancy remains a valuable tool for summarizing post-pandemic population mortality trends. Compared to excess mortality, life expectancy deficits give more weight to deaths at younger ages, and are age-standardized to allow comparisons across countries with different population sizes and age structures.

Several limitations merit consideration. First, our counterfactual estimates rely on extrapolative Lee-Carter forecasts, which assume continuation of pre-pandemic mortality trends. If countries would have over- or under-performed these projections even in the absence of COVID-19, our estimates may partly reflect deviations unrelated to the pandemic. For example, while traditional Lee-Carter models did well overall in forecasting US mortality trends since 2000, they did not anticipate the significant slowdown in improvements since 2014 (Basellini et al. 2023). Djenunde et al. show that mortality improvements were worse than forecasted after 2010 for many high-income countries (Djeundje et al. 2022). Thus, it is possible that the persistent deficits observed in countries such as the Netherlands may partially reflect slowdowns in mortality improvements that would have occurred regardless of COVID but were not reflected in their Lee-Carter estimates. Conversely, countries that would have seen faster-than-expected mortality improvements even in the absence of COVID-19 may appear to have better “recoveries.” For countries with stagnating or slow mortality improvements prior to 2020, such as the USA and UK, returning to their pre-pandemic trajectory reflects a less significant achievement than for countries that were seeing rapid improvements prior to the pandemic (such as Eastern Europe). Overall, the farther we get from pre-pandemic mortality data, the harder it will be to calculate a valid counterfactual and to separate any ongoing COVID-19 impacts from other mortality dynamics. Nevertheless, the persistence and cross-country consistency of deficits through 2024 and the robustness of our estimates to different assumptions including a shorter, 10-year baseline window suggests that forecast uncertainty alone is unlikely to explain the observed patterns.

Second, our country-level analysis does not capture the substantial within-country variation in mortality trajectories since 2020. For example, there were significant socioeconomic and race/ethnic differences in COVID mortality in the US (Aburto et al. 2022, Case and Deaton 2024) and globally (McGowan and Bambra 2022; Subedi et al. 2025). These differential mortality shocks likely imply distinct recovery trajectories across sociodemographic groups, which are obscured by national averages. Within-country research on the evolution of health inequalities since the pandemic, and specifically which groups may be lagging in mortality recovery, will be important going forward.

Finally, our analysis is also limited to the countries with available age-specific mortality data for the period covered and thus doesn’t capture the full range of COVID-19 mortality experiences globally. Data from Mexico through 2022 suggest a pattern consistent with our “second wave peak” category, with large life expectancy losses in 2020 and 2021 followed by a sharp rebound in 2022 (Zazueta-Borboa et al. 2025). Analysis from Australia and Hong Kong shows a pattern of delayed and milder COVID mortality consistent with our “late deficit” category, with life expectancy improving in 2020 and not falling until 2022, but with overall positive gains from 2020-2022 (Adair et al. 2023; Law et al. 2025). China maintained stringent zero-COVID restrictions until late 2022, followed by a rapid rate of population infection over a short time. While reliable mortality data from China was not available to include in our analysis, some modelled estimates from 2023 suggest that China’s death rate was considerably higher than other countries that delayed infection like Japan, but lower than countries that saw elevated mortality throughout the pandemic like the USA (Du et al. 2023; Fung et al. 2026). As data from more countries becomes available, it will be important to examine the full global scope of COVID-19 mortality experiences and recovery.

### Conclusion

Five years after the emergence of SARS-CoV-2, the pandemic has proved to be more than a brief interruption to steady mortality improvements. The variation in mortality experiences across countries highlights that the pandemic’s impact was shaped by a variety of factors, including the timing of initial spread, pre-existing population health vulnerabilities, and broader social and institutional responses. This could reflect both the virus’s direct biological impacts, disruptions to health care systems, and other downstream social repercussions of the pandemic. Continued monitoring of mortality trends by age, cause of death, and sociodemographic subgroups will be essential to understand whether the impact of COVID-19 continues to shape population health and mortality trajectories for years to come.

## Data Availability

Reproducible data and code for all analysis are available at: https://doi.org/10.5281/zenodo.21808782

## Acknowledgements

This paper was presented at the 2025 Annual Meeting of the Population Association of America. We thank participants at the Berkeley Brown Bag Seminar and the Human Mortality Database Symposium for their helpful feedback. Special thanks to Linus Schöley, who pointed out that the flags of Luxemburg and Netherlands are too easy to mix up without further clues.

## Author contributions

Conceptualization: all authors; data preparation: J.S, A.P., H.L.; data analysis: J.S.; interpretation: all authors; manuscript draft: J.B.D.; critical manuscript revision: all authors

## Supplementary Material

**Table S1.**
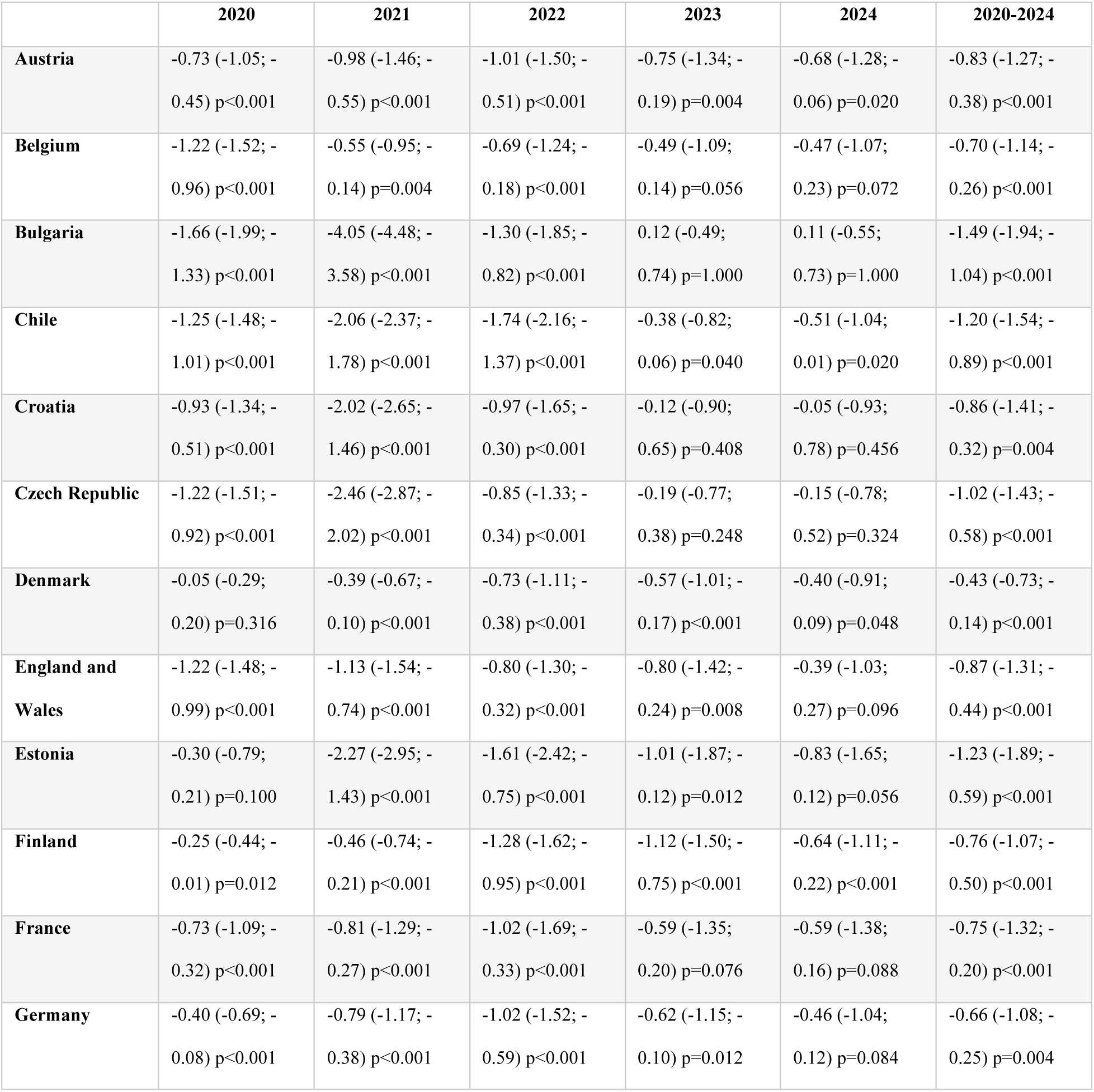
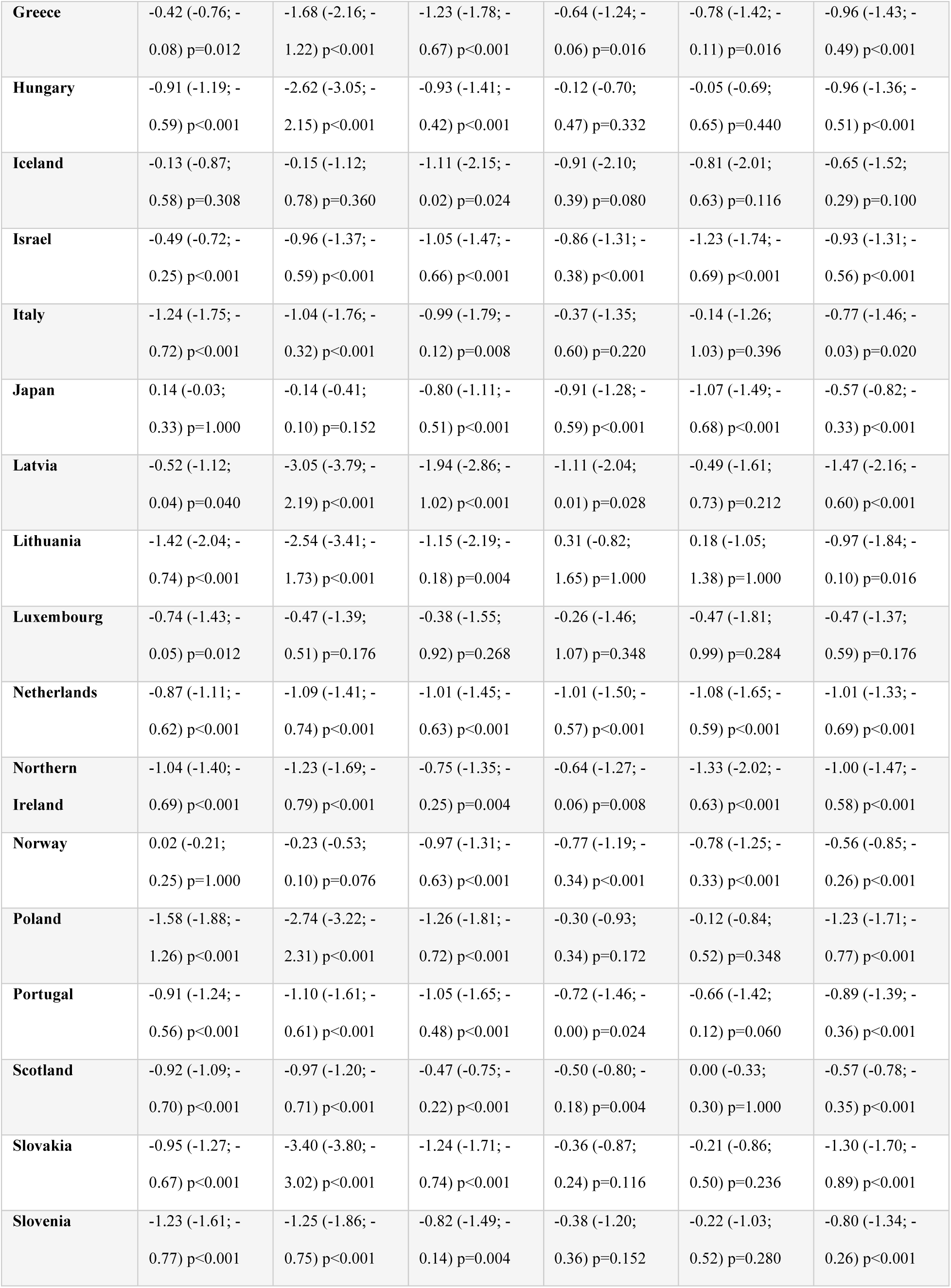
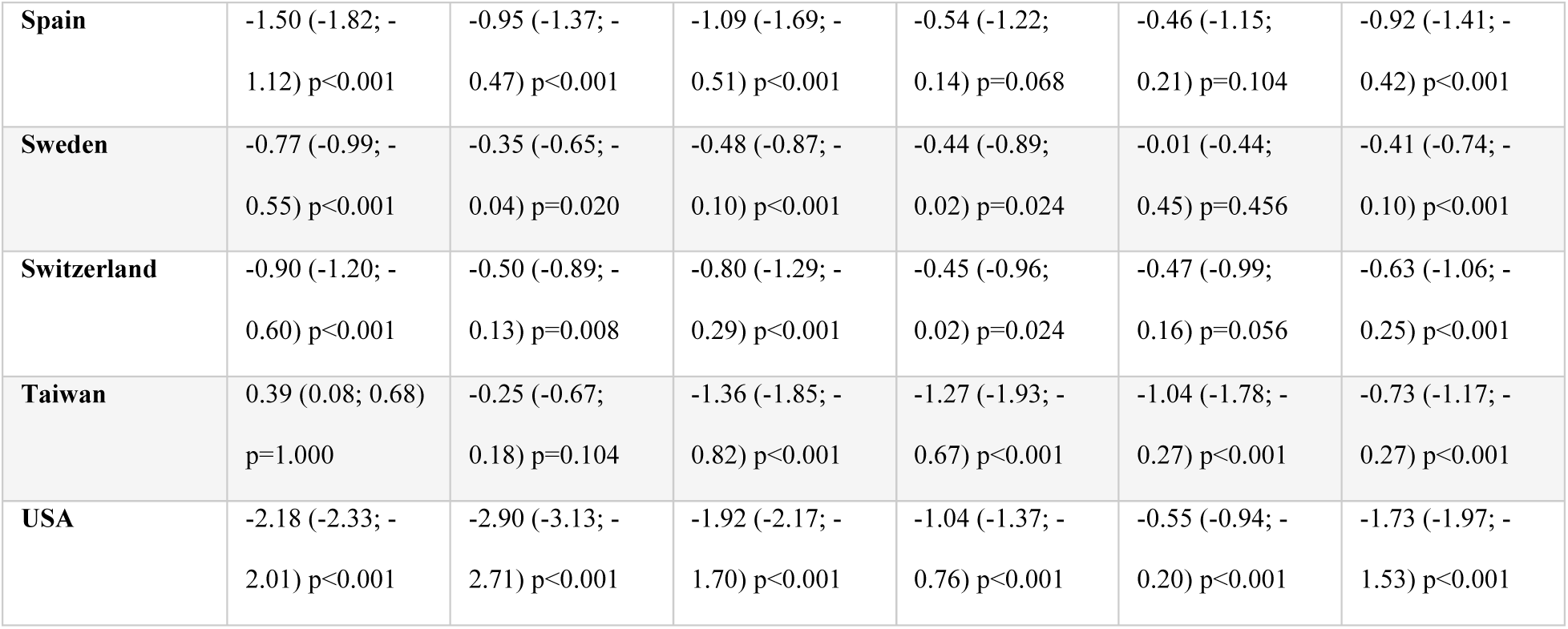
Annual and overall life expectancy deficits 2020 through 2024 for both sexes combined. Shown are the differences between actual and projected life expectancy for the years 2020 through 2024 across 34 high-income countries. 95% prediction intervals are given in parenthesis. The p-value gives the probability of observing the given life expectancy deficit or higher had pre-pandemic mortality trends continued.

**Figure S1.**
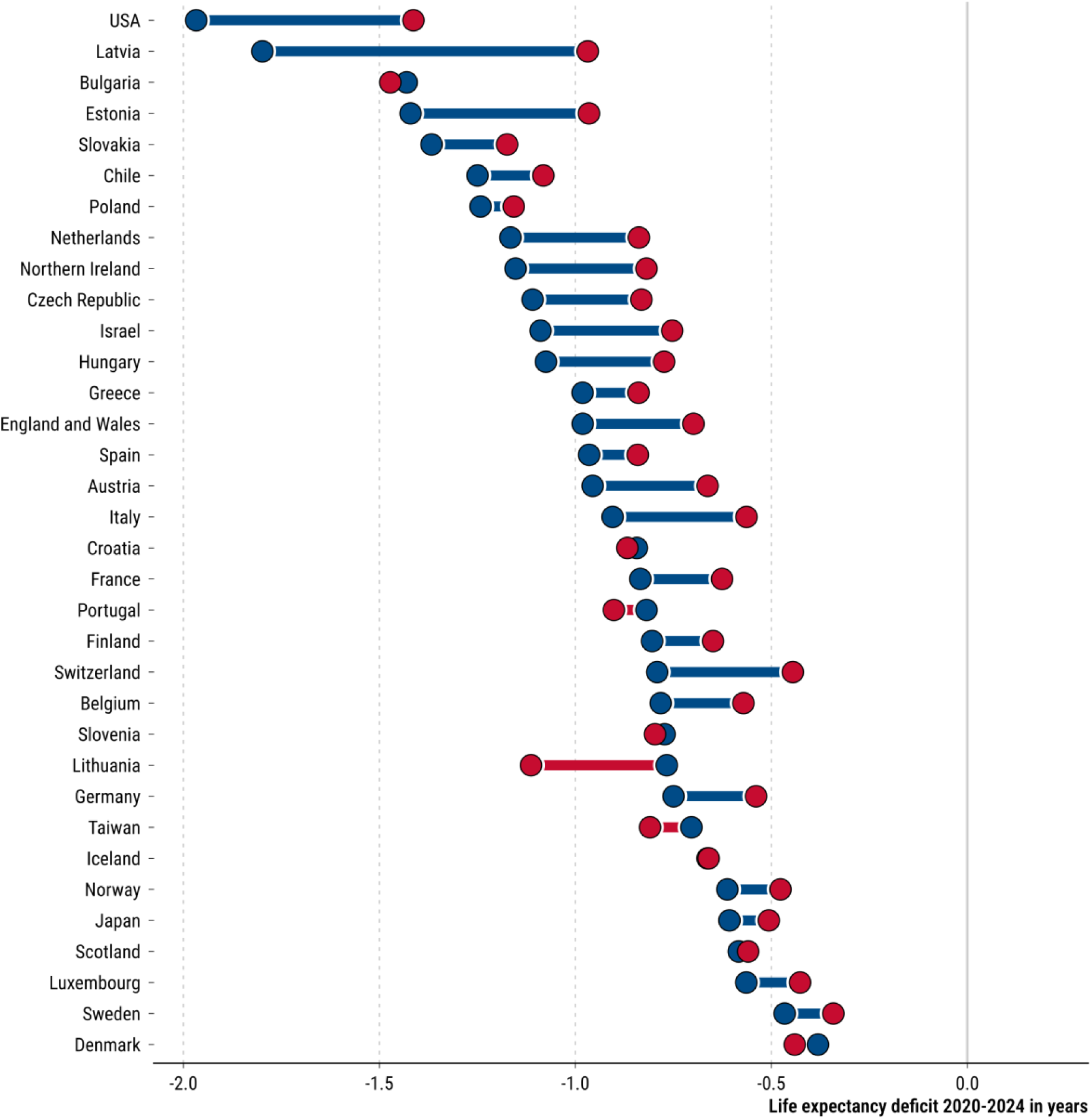
Life expectancy deficit for 2020-2024 overall, by sex. The red dots refer to female, the blue dots to male deficits. Correspondingly, the colour of the connecting line indicates either excess female or excess male deficit.

**Figure S2.**
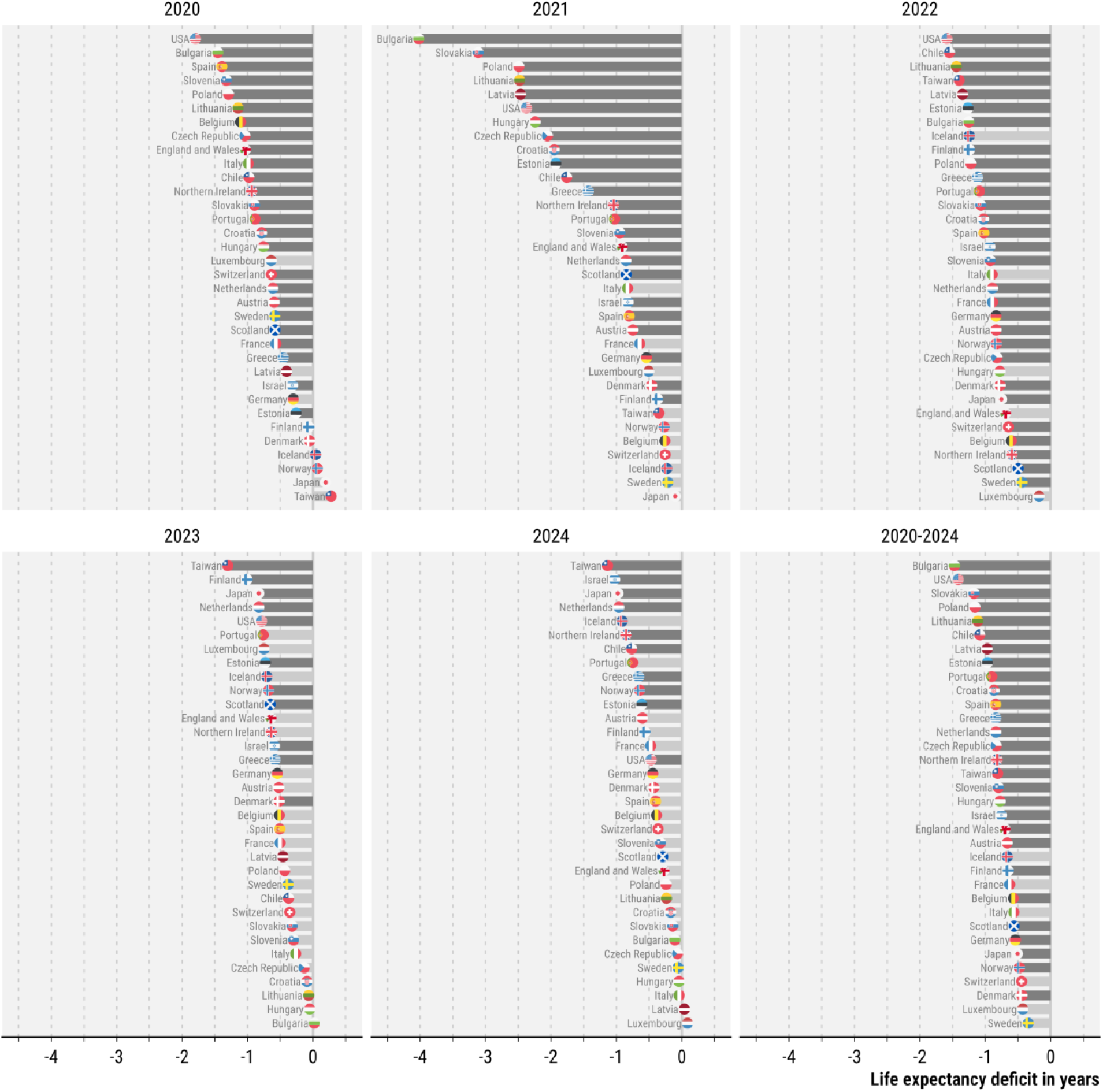
Country rankings of female life expectancy deficits for each year in the interval 2020-2024 and in total. Shown are the sex differences in life expectancy deficits for 34 countries. A dark grey bar indicates a p-value of ≤0.05 for the probability that this deficit would be observed under the continuation of pre-pandemic trends.

**Figure S3.**
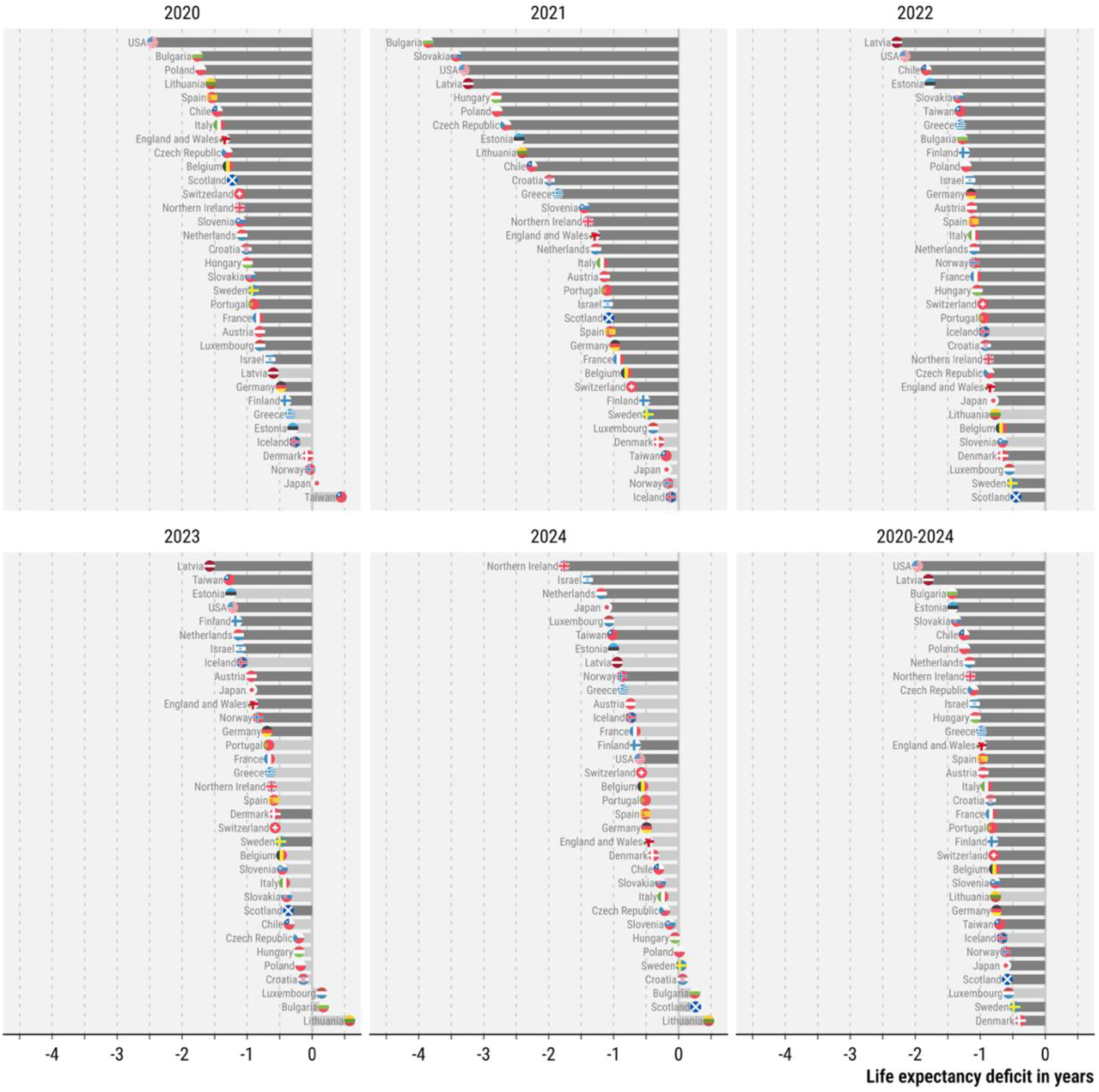
Country rankings of male life expectancy deficits for each year in the interval 2020-2024 and in total. Shown are the sex differences in life expectancy deficits for 34 countries. A dark grey bar indicates a p-value of ≤0.05 for the probability that this deficit would be observed under the continuation of pre-pandemic trends.

**Figure S4.**
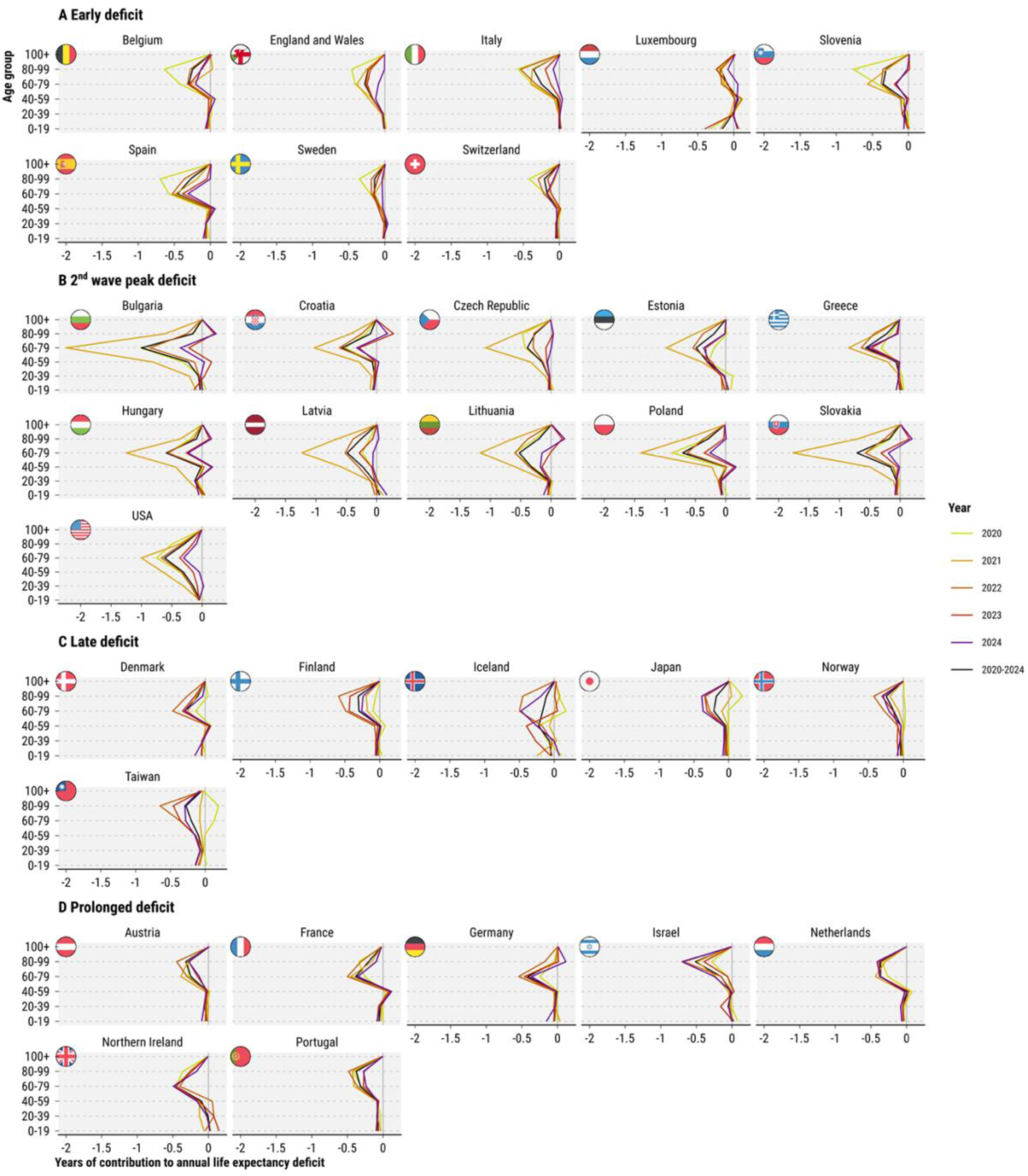
Age specific contributions to female annual life expectancy deficits 2020 through 2024 and for all years combined.

**Figure S5.**
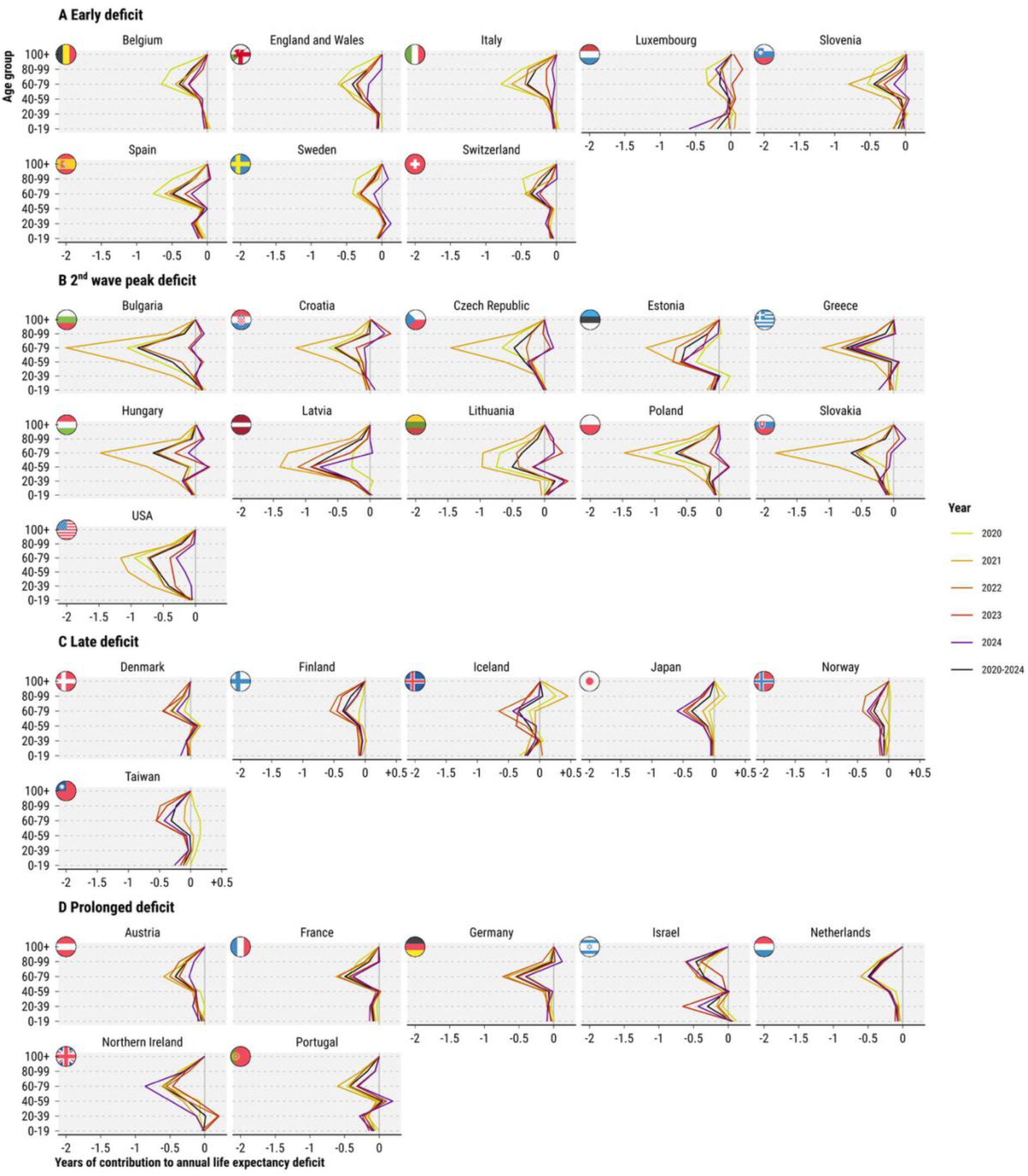
Age specific contributions to male annual life expectancy deficits 2020 through 2024 and for all years combined.

**Figure S6.**
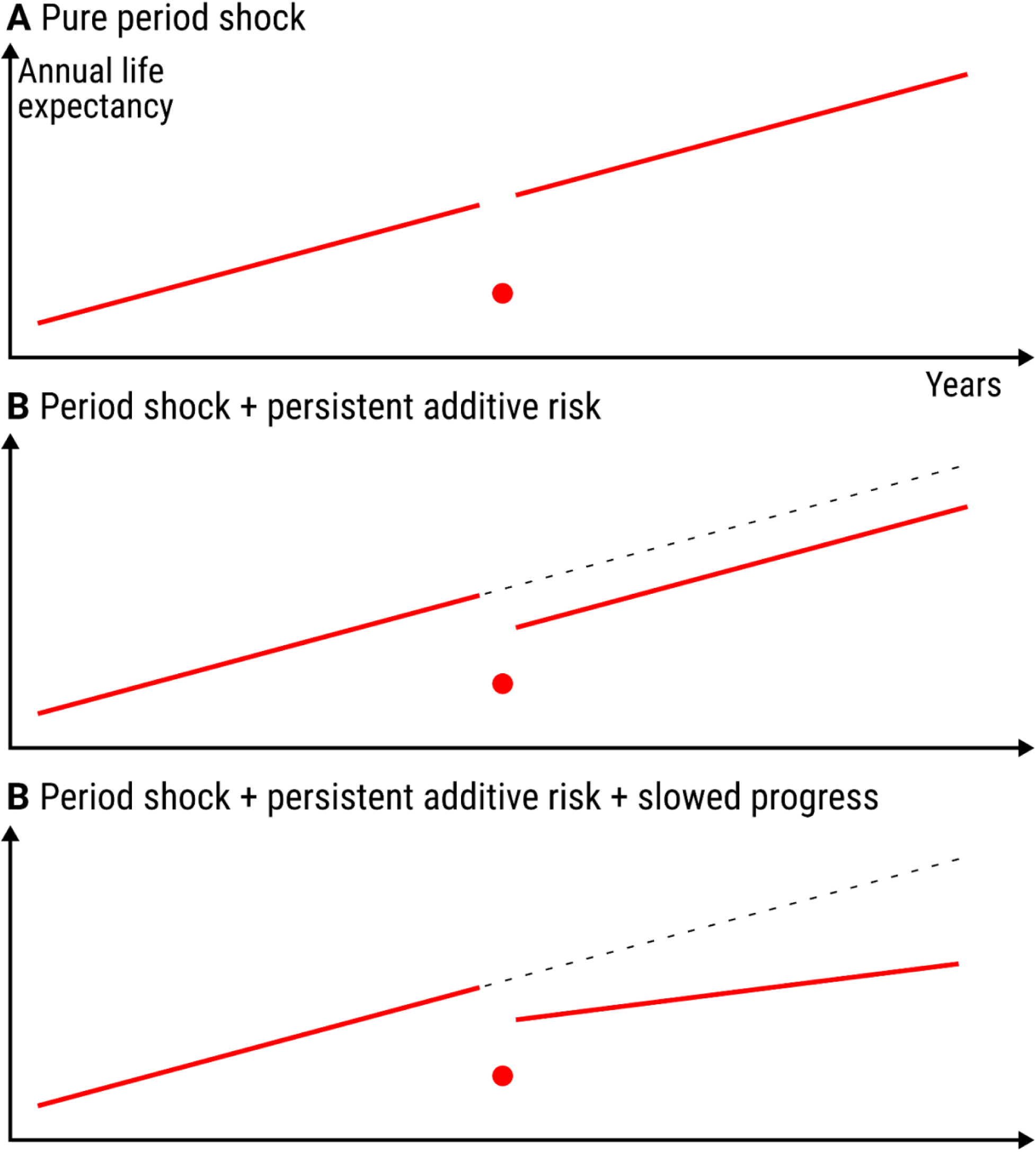
Stylized scenarios of mortality shocks on life expectancy trajectories.

## 1. Summary

We calculate life tables and associated life expectancies by sex for a selection of 34 countries for the years 2000 through 2024. For the years 2020 through 2024 we calculate counterfactual life expectancies based on the continuation of pre-pandemic mortality trends and derive associated life expectancy deficits – the difference between actual and counterfactual life expectancy – as an age-standardized and trend-adjusted measure of excess mortality.

The data basis for our calculation are midyear population and annual death counts by age, sex, and country sourced from the Short Term Mortality Fluctuations Database (HMD-STMF [1]), the Human Mortality Database (HMD, [2, 3]), the UN World Population Prospects (WPP-2024 [4]), and national statistical offices (ONS [5], CDC [6]). For a complete list of sources by country see methods appendix “Data sources”. We selected all countries represented in the Human Mortality Database for which we could find reliable death counts by age and sex for 2024 as of February 5, 2026, the date on which we froze our source data, either in the HMD’s annual data series, or in HMD-STMF. Shared characteristics of all included countries are a) that they are designated as “high-income” by the World Bank’s income classification, and b) that their data series have a high degree of reliability and comparability which is ensured by the ongoing documentation, correction, and harmonization work by the Human Mortality Database’s team.

Age-specific death counts are harmonized to a single-year age grouping with open age group 100+ using the PCLM method [7, 8]. Weekly death counts are aggregated to annual counts and midyear population estimates are converted to person-years of exposure adjusting for the effect of leap weeks. To adjust for systematic bias in registered deaths between annualized STMF data and HMD annual totals we estimate correction factors based on a statistical model (see methods appendix “Bias correction of age-specific annual death counts”).

Given the harmonized annual death counts and population exposures by age and sex we calculate period life tables using the standard piecewise-exponential model. We then fit a Poisson-Lee-Carter model [9] using the R library StMoMo [10] to the available pre-pandemic data since 2000 and forecast counterfactual age-specific mortality rates and death counts over the years 2020 through 2024. Life expectancy deficits (LED) are then derived along with P-scores [11] and the mean unfulfilled lifespan (MUL, [12]) as alternative measures of excess. We decompose the life expectancy deficits into age-specific contributions using the Arriaga method [13]. The clustering of life expectancy deficits into four groups is achieved via a hierarchical clustering algorithm implemented in the R package dtwclust [14] which we run across the country-specific trajectories (see methods appendix “Clustering of life expectancy deficits” for details).

Throughout the analysis population exposures and annual death counts by sex are aggregated as needed to derive results for both sexes combined and for the joint period 2020-2024.

We perform simulation-based uncertainty propagation throughout our analysis via a Poisson parametric bootstrap of the life table estimates and a random walk simulation of the Lee-Carter forecast. Based on this we derive a p-value for the estimated life expectancy deficit, i.e. the probability of observing or exceeding an estimated life expectancy deficit given the continuation of pre-pandemic trends (see methods appendix “Life table inference” for details).

We ensured that our findings are robust against changes in counterfactual, changes in fitting period, and changes in clustering procedure (see methods appendix “Robustness and sensitivity”). The complete analysis workflow is explained in detail in the methods appendix. The analysis script is available from https://github.com/jschoeley/e0deficit.

## 2. Data

### 2.1 Data sources

The inputs for our analysis are death and population counts by age, year, sex, and country. We source^1^ the death counts from:

- The Human Mortality Database (HMD, [2, 3]) for annual death counts and exposures by age and sex for those countries for which a complete series is available through 2024.
- The Short Term Mortality Fluctuations Database (HMD-STMF, [1]), an input database for weekly death counts by age and sex across countries. The data are updated weekly and include the most recent information on death counts. We derive annual counts from this source in cases where the HMD does not provide data through 2024.
- The UK’s Office for National Statistics (ONS, [5]) annual death counts by age and sex for England and Wales. We use this data for years 2010 through 2019 because the age grouping in the HMD-STMF data for these years is too coarse for reliable life expectancy estimates.
- The US Centers for Disease Control and Prevention (CDC, [6]) for annual death counts by age and sex for the United States. We use this data for years 2000 through 2019 for the US, lacking any HMD-STMF data for these years.

Population data form the basis for our person-year exposure estimates and are sourced from:

- The World Population Prospects 2024 (WPP-2024, [4]) for midyear population estimates and projections by age, year, and country.
- The HMD for midyear population estimates by age, year, and country for those countries for which a complete series is available through 2024 and for the regions of the UK.

For a list of data sources by country and year see Table 1.

We selected all countries represented in the Human Mortality Database for which we could find reliable death counts by age and sex for 2024 as of February 5, 2026, the date on which we froze our source data, either in the HMD’s annual data series, or in HMD-STMF.

Countries which were excluded under these criteria are Australia (no 2024 annuals, coarse age grouping in STMF), Canada (no 2024 annuals, coarse age grouping in STMF), South Korea (no 2024 annuals, coarse age grouping in STMF), New Zealand (no 2024 annuals, coarse age grouping in STMF), Hong Kong (no 2024 annuals or STMF), Russia (no data since 2022), and Ukraine (no data since 2014).

Shared characteristics of all included countries are a) that they are designated as “high-income” by the World Bank’s income classification, and b) that their data series have a high degree of reliability and comparability which is ensured by the ongoing documentation, correction, and harmonization work by the Human Mortality Database’s team.

We excluded Scotland and Chile from the age decomposition as we found artifacts in the age patterns of mortality in both countries. For both countries we relied on the preliminary STMF data. These artifacts are localized in very old age and adolescent mortality and therefore have little influence on life expectancy estimates but some influence on the age decomposition.

**Table 1:** Data sources by country and year.

| Country | Period | Death counts | Population |
| --- | --- | --- | --- |
| Austria | 2000-2024 | HMD-STMf | WPP-2024 |
| Belgium | 2000-2024 | HMD | HMD-EST |
| Bulgaria | 2000-2024 | HMD-STMf | WPP-2024 |
| Switzerland | 2000-2024 | HMD | HMD-EST |
| Chile | 2000-2015 | HMD | WPP-2024 |
| Chile | 2016-2024 | HMD-STMf | WPP-2024 |
| Czech Republic | 2000-2004 | - | WPP-2024 |
| Czech Republic | 2005-2024 | HMD-STMf | WPP-2024 |
| Germany | 2000-2024 | HMD-STMf | WPP-2024 |
| Denmark | 2000-2024 | HMD | HMD-EST |
| Estonia | 2000-2024 | HMD | HMD-EST |
| Spain | 2000-2024 | HMD-STMf | WPP-2024 |
| Finland | 2000-2024 | HMD | HMD-EST |
| France | 2000-2024 | HMD-STMf | WPP-2024 |
| England and Wales | 2000-2009 | - | HMD-EST |
| England and Wales | 2010-2019 | ONS | HMD-EST |
| England and Wales | 2020-2022 | HMD-STMf | HMD-EST |
| England and Wales | 2023-2024 | HMD-STMf | HMD-PRJ |
| Northern Ireland | 2000-2021 | HMD | HMD-EST |
| Northern Ireland | 2022 | HMD-STMf | HMD-EST |
| Northern Ireland | 2023-2024 | HMD-STMf | HMD-PRJ |
| Scotland | 2000-2022 | HMD-STMf | HMD-EST |
| Scotland | 2023-2024 | HMD-STMf | HMD-PRJ |
| Greece | 2000-2014 | HMD | WPP-2024 |
| Greece | 2015-2024 | HMD-STMf | WPP-2024 |
| Croatia | 2000 | - | WPP-2024 |
| Croatia | 2001-2024 | HMD-STMf | WPP-2024 |
| Hungary | 2000-2024 | HMD-STMf | WPP-2024 |
| Israel | 2000-2024 | HMD-STMf | WPP-2024 |
| Iceland | 2000-2024 | HMD-STMf | WPP-2024 |
| Italy | 2000-2010 | - | WPP-2024 |
| Italy | 2011-2024 | HMD-STMf | WPP-2024 |
| Japan | 2000-2024 | HMD | HMD-EST |
| Lithuania | 2000-2024 | HMD | HMD-EST |
| Luxembourg | 2000-2024 | HMD-STMf | WPP-2024 |
| Latvia | 2000-2024 | HMD | HMD-EST |
| Netherlands | 2000-2024 | HMD-STMf | WPP-2024 |
| Norway | 2000-2024 | HMD | HMD-EST |
| Poland | 2000-2024 | HMD-STMf | WPP-2024 |
| Portugal | 2000-2024 | HMD | HMD-EST |
| Sweden | 2000-2024 | HMD | HMD-EST |
| Slovenia | 2000-2024 | HMD-STMf | WPP-2024 |
| Slovakia | 2000-2024 | HMD | HMD-EST |
| Taiwan | 2000-2024 | HMD-STMf | WPP-2024 |
| USA | 2000-2019 | CDC | WPP-2024 |
| USA | 2020-2024 | HMD-STMf | WPP-2024 |

### 2.2 Data harmonization

#### 2.2.1 Death count age harmonization

As the HMD-STMF input data are not harmonized in terms of age grouping we perform^2^ a model-based ungrouping of deaths into single ages for each region-year-sex stratum using the Penalized Composite Link Method (PCLM) [7] implemented in the R package ungroup [8].

The original death counts over single-year age groups **d**^single^ =(*d _j_*_=1_,…, *d_J_*)^⊺^ have been grouped (summed) into discrete age bins **d**^grouped^ = (*d_i_*_=1_,…, *d_I_*)^⊺^of varying width. We assume the grouped death counts to arise from a Poisson distribution with mean *μ_i_*:

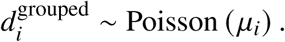

The PCLM method seeks to recover the means of the original single-year death counts, ***γ*** = *(γ_j_*_=1_,…, *γ_J_*)^⊺^. To that end, one can encode the age aggregation into a binary composition matrix **C** with dimensions *I* ×*J* such that **d**^grouped^ = **Cd**^single^. The grouped Poisson rates are then connected to the single age means via the composite link

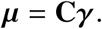

By specifying that the ***γ*** are a smooth function of age,

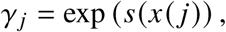

where *s*(*x*(*j*)) is a P-spline over the starting age *x* of single-year age groups *j* ∈ 1,…, *J*, a penalized likelihood solution can be found for the vector of mean death counts by single age. To get an integer estimate of the death count in single age *x* we round *γ_x_* (*j*) down to the next integer,

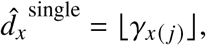

such that *d̂_x_* corresponds to the mode of the Poisson distribution with rate *γ_x_*_(_ *_j_*_)_.

Because the age grouping scheme may vary within a year, the ungrouping is separately applied to each region × sex × year × age pattern combination in the input data. In a final step, the harmonized sub-annual HMD-STMF death counts are summed into annual death counts by age, *d̂_x__y_*.

#### 2.2.2 Exposure time calculation

We derive person-years of exposure by age and year, *E_xy_*, from the midyear population estimates, 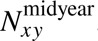. We choose the HMD midyear population for exposure calculation for those countries where a complete series is available through 2024 and for the regions of the UK. Otherwise we choose the World Population Prospects 2024 data, which are always complete throughout 2024. To complete HMD exposures for the UK regions we perform a stable population projection based on the most recent HMD population data and 5-year average (2015 through 2019) age-specific mortality and fertility rates. Exposures are adjusted for the length of the observation period within a year, taking into account weeks with missing data and leap weeks.

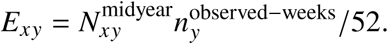

#### 2.2.3 Bias correction of age-specific annual death counts

The purpose of our bias correction model is to correct any systematic deviations between our age-specific mortality rate estimates and the official annual estimates. While the HMD-STMF death counts allow for timely estimates of current mortality they sometimes deviate substantially from official annual estimates due to different population definitions and reporting guidelines in the weekly death reporting compared to the annual reporting. Because these systematic biases are stable in time, they can be estimated and corrected. To that end we estimate^3^ a correction factor *Ĉ_xy_* which, if multiplied with the single-age annual death counts, *d̂_xy_*, yields the corrected counts:

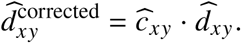

We estimate the correction factor via the Poisson model

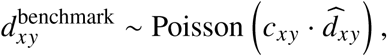

where the benchmark death counts are taken from the HMD and

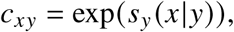

is a smooth age effect estimated separately for each year capturing the time-varying deviations between unadjusted and benchmark death counts. The correction factor from the last year with available benchmark data will be used in subsequent years.

Figures 1 and 2 show the estimated correction factors and the bias-corrected age-specific death counts by year for the example of Austrian females. The model is fit separately to each region and sex.

**Figure 1:**
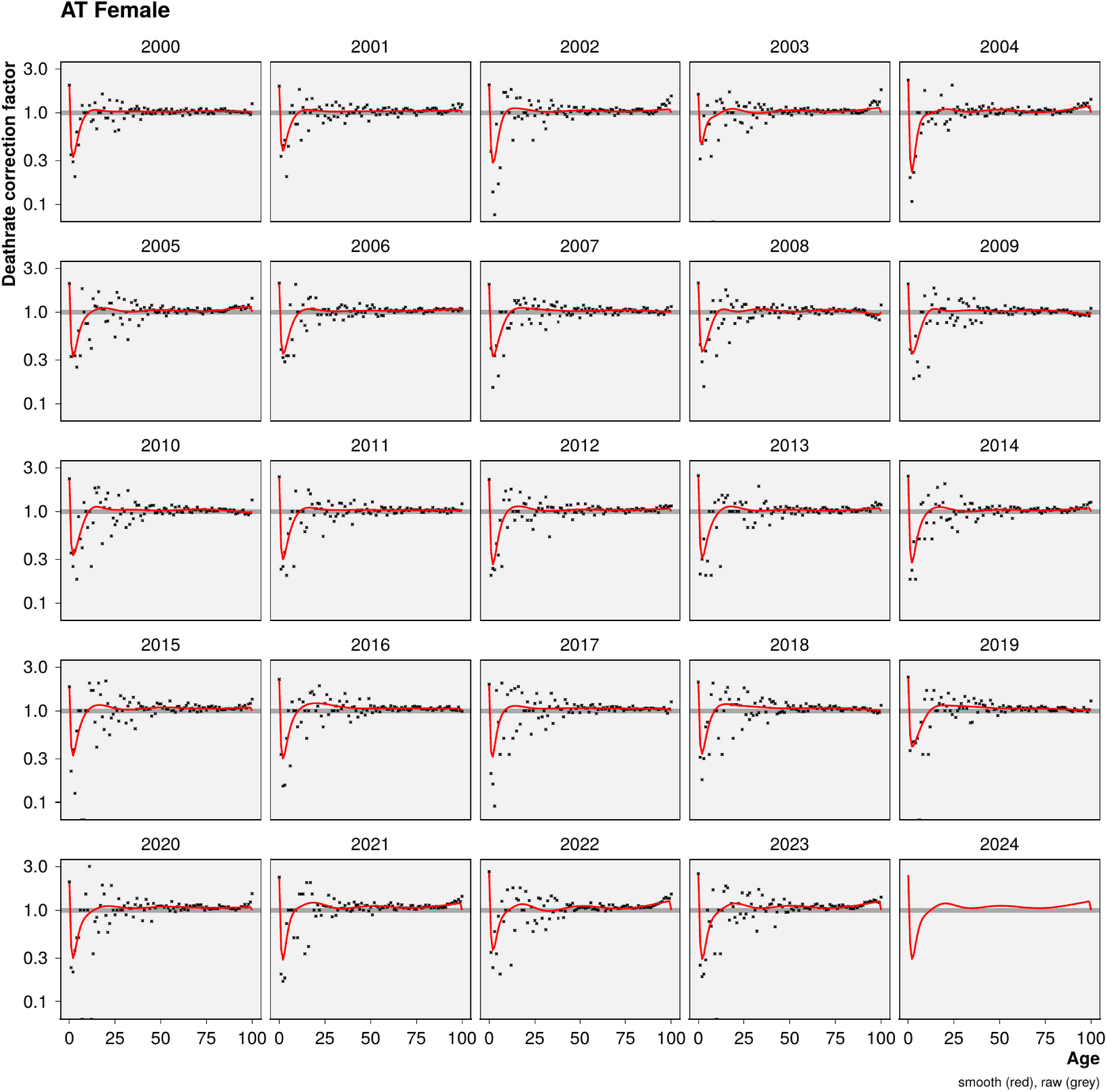
Demonstration of estimated (red) bias correction factors *c_xy_* vs. raw correction factors (black) for Austrian females.

**Figure 2:**
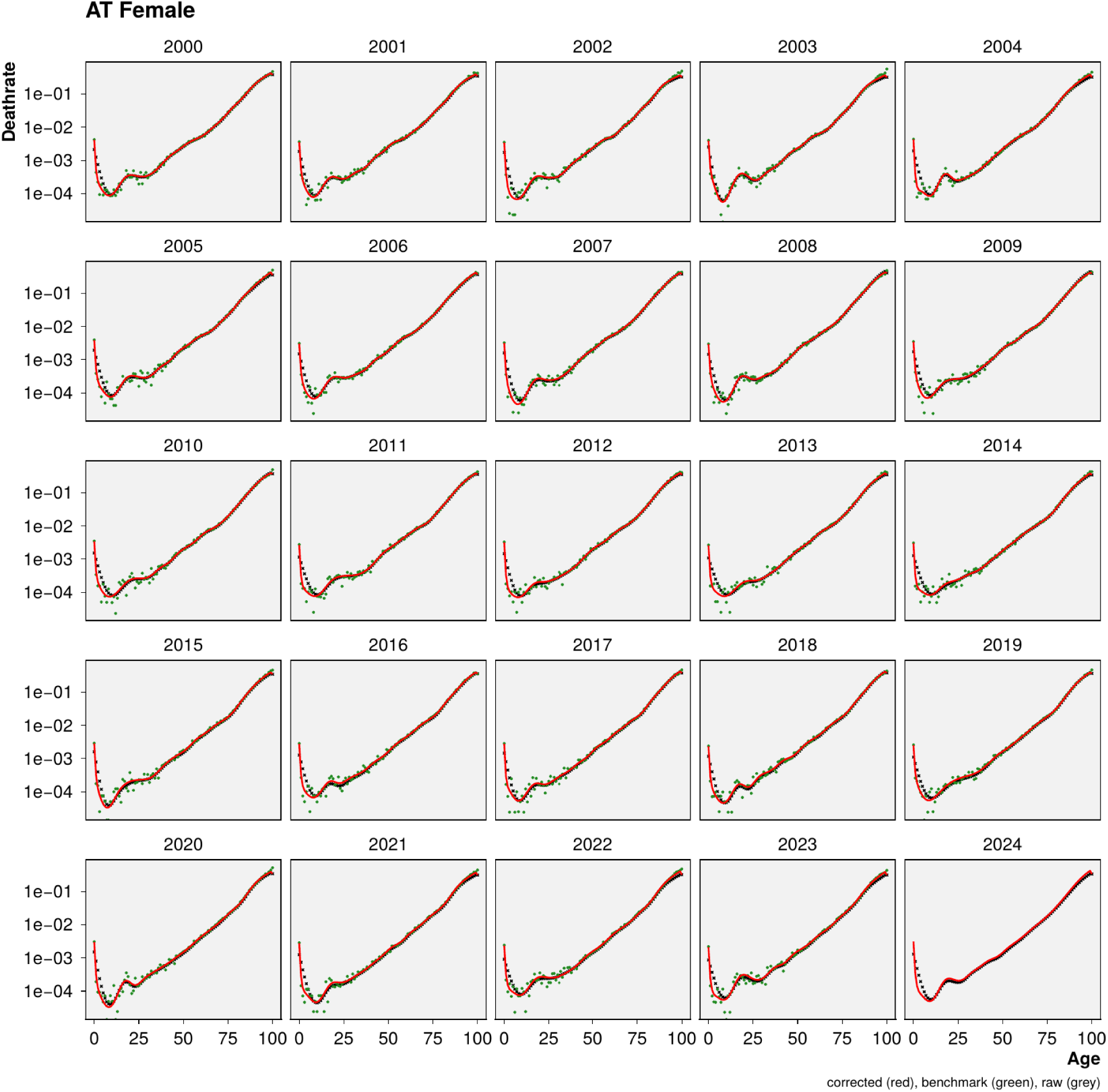
Demonstration of bias-corrected ungrouped age-specific death rates (red) vs. unadjusted death rates (black) and HMD benchmark death rates (green) for Austrian females.

## 3. Analysis

### 3.1 Counterfactual mortality forecast

In order to calculate the life expectancy under continuation of pre-pandemic trends we forecast^4^ age-specific mortality rates over the years 2020-2024 using a Poisson-Lee-Carter model given the data available up to January 2020.

We fit a Poisson-Lee-Carter model [9] where death counts at single age *x* and single year *y* are distributed

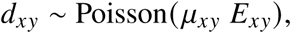

with death rates

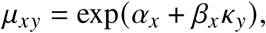

and identifiability constraints Σ*_y_ κ_y_* = 0 and Σ*_x_*β*_x_*=1.

We forecast *μ_xy_* past the fitting period via the usual random walk with drift over *κ_y_*,

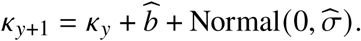

We estimate the variance and mean difference of the estimated year-on-year *κ_y_* changes and simulate a corresponding random walk with drift over the years 2020-2024. The resulting forecasts of *κ* are then used to derive a corresponding surface of age-period mortality rates 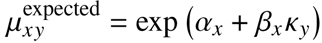 for forecast horizon *y* ∈ {2020, 2021,…, 2024}. We fit the model using the R library StMoMo [10]. Forecast uncertainty is propagated throughout the rest of the analysis via 250 samples of the forecasted 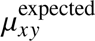.

### 3.2 Life table inference

We perform a parametric bootstrap of our life table estimates^5^ based on 250 Poisson replicates of actual and expected age-specific death counts for any population under consideration (for brevity sex, country, and year indices are omitted in this section).

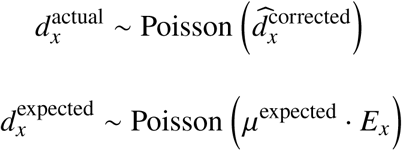

At this point we aggregate the simulated death counts along with the person-years of exposure into larger populations of interest, e.g. male and female are summed to total population and annual data are summed to multi-year aggregates. All of the following analysis steps are also performed for the newly created aggregated populations.

For any given population and for each sample draw we calculate the actual and expected central death rates by age as

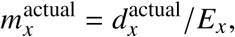

and

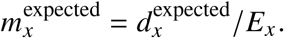

These rates are then transformed into life tables yielding actual and expected life expectancies, 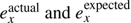, for each population, year, and simulation draw. We write *n_x_* = 1 for *x* ≠ *ω* and *n_ω_* = ∞ for the width of the age group starting at *x* with *ω* = 100 being the open age group closing the life table. Under the piecewise constant assumption we calculate the life tables as

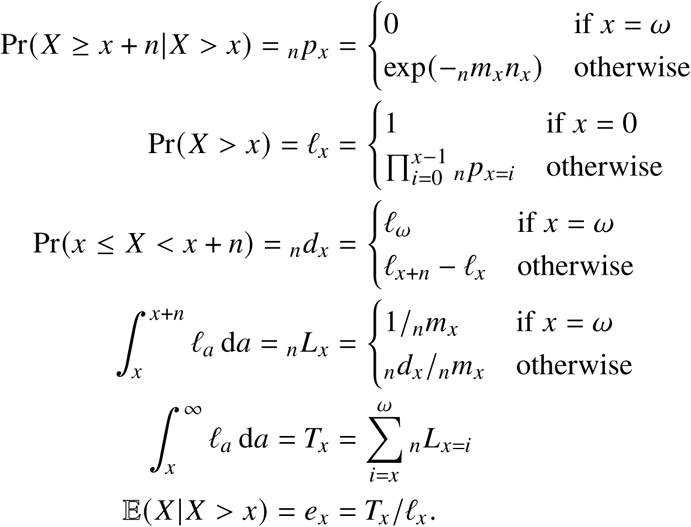

Now the *life expectancy deficit* can be calculated as the difference between actual and counterfactual life expectancy, the baseline life expectancy under the continuation of pre-pandemic trends,

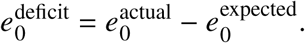

Note that we define the deficit such that it is negative if actual life expectancy falls short of the expectation, i.e. if there are excess deaths, and positive if there is a life expectancy surplus, i.e. if the expectation is outperformed.

Life expectancy deficits do not necessarily indicate a structural deviation from expected mortality trends as both the expected and the observed life tables are subject to uncertainty. Uncertainty in the expectation is mainly due to forecasting error – we do not know exactly how age-specific mortality rates would have developed in the absence of the pandemic. This uncertainty is captured by the random walk of the forecasting model described earlier. Additionally, aleatoric uncertainty in the death counts induces uncertainty in the estimated central death rates which is reflected via our Poisson resampling scheme.

Due to the stated sources of randomness one may observe life expectancy deficits even under the continuation of pre-pandemic trends. Thus we perform inference about the estimated *e*^deficit^ by calculating a p-value stating the probability of estimating a life expectancy deficit as large as the realized deficit under the continuation of pre-pandemic trends. Life expectancy deficits under the null hypothesis of pre-pandemic trends are defined as

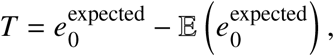

and we derive the sampling distribution of *T* from our sample draws of 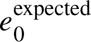. We can then calculate the probability of our calculated 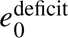 given pre-pandemic trends as

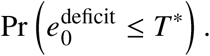

Figure 3 demonstrates the procedure.

**Figure 3:**
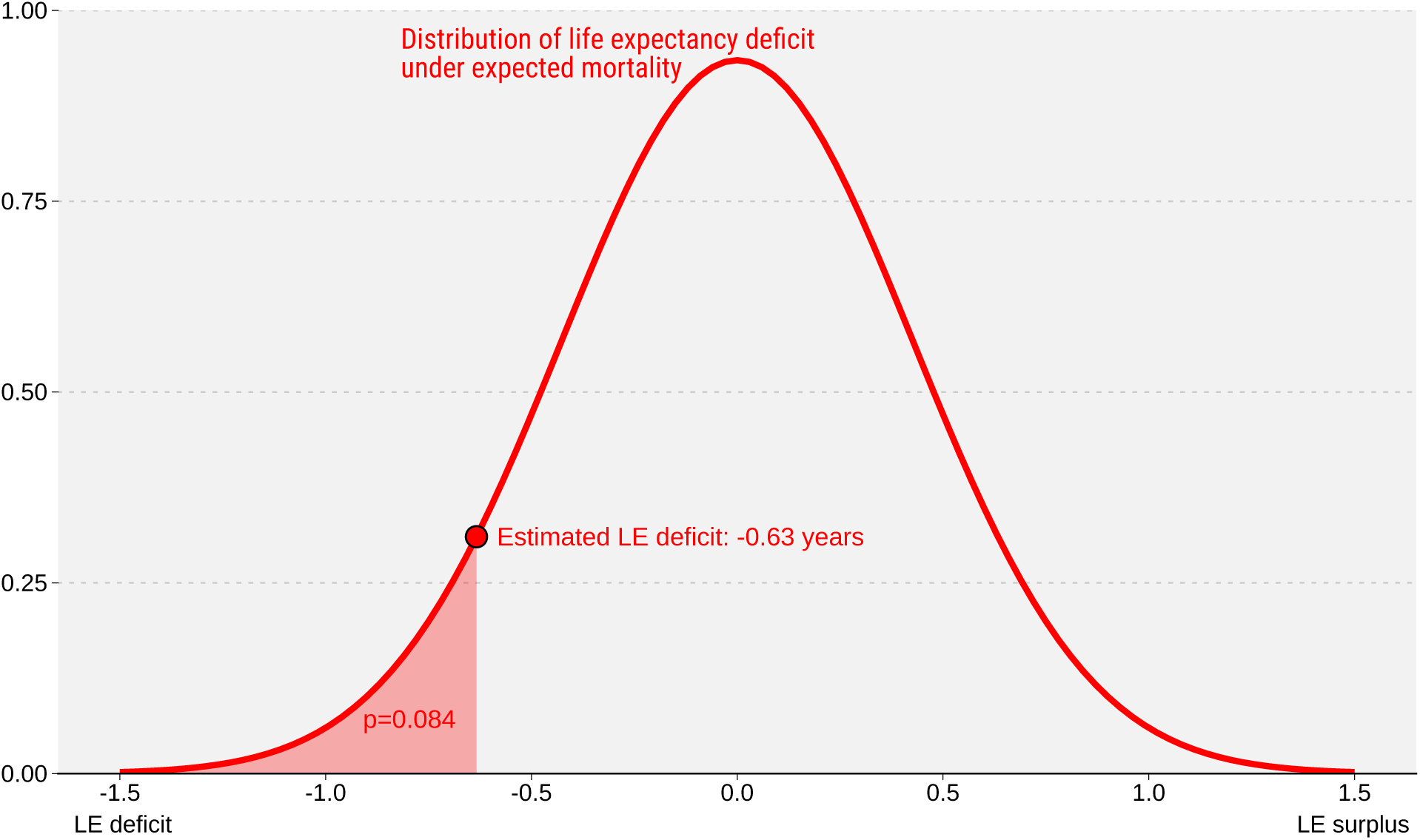
Demonstration of p-value calculation for estimated life expectancy deficit.

### 3.3 Clustering of life expectancy deficits

We calculate time series of life expectancy deficits, 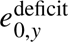, for years 2020 through 2024 by region. These series are then clustered^6^ into four groups: A) Early deficit (peak deficit in 2020 or 2021 without pronounced worsening in 2021), B) Second wave peak deficit (pronounced peak deficit in 2021), C) Late deficit (worsening of deficits starting 2022), and D) Prolonged deficit (deficits since 2020 without pronounced peak or trend).

The group assignment for each country’s deficit trajectory is formalized by employing hierarchical clustering with a pre-specified cluster number of four. We cluster on deficit series for both sexes combined. Prior to clustering we derive three time series features for each region *c*: the range of the deficits, 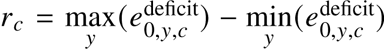, the centered and range-normalized trajectory of deficits, 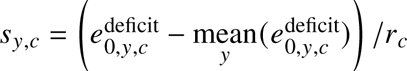, and the first differences of *s_y_*_,*c*_ over time. We then run the hierarchical clustering algorithm implemented in the R package dtwclust [14] with Euclidean distance and Ward’s minimum variance method on the combined vector of features across regions to determine each region’s cluster. The resulting clustering captures the phenomenology of the four qualitative groups described above.

### 3.4 Age decomposition analysis

We perform the Arriaga decomposition [13] to attribute life expectancy deficits to age-specific excess mortality^7^. As the Arriaga decomposition is analytic and can be expressed as a series of simple arithmetic operations it lends itself well to simulation-based inference where computation speed is vital. Based on the previously estimated life table quantities we calculate

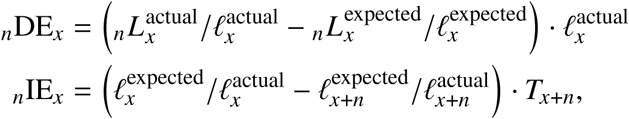

which yields the contribution of excess mortality at age *x* to the overall life expectancy deficit:

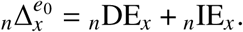

## 4. Robustness and sensitivity

### 4.1 Benchmark tests

We compare^8^ our life expectancy estimates to available estimates by Eurostat and the Human Mortality Database. As intended, our estimates are closely aligned with the HMD reported life expectancies. In some regions, Spain for example, small discrepancies exist between HMD’s and Eurostat’s estimates. However, the overall shape of the life expectancy trajectories is qualitatively the same under either set of estimates.

**Figure 4:**
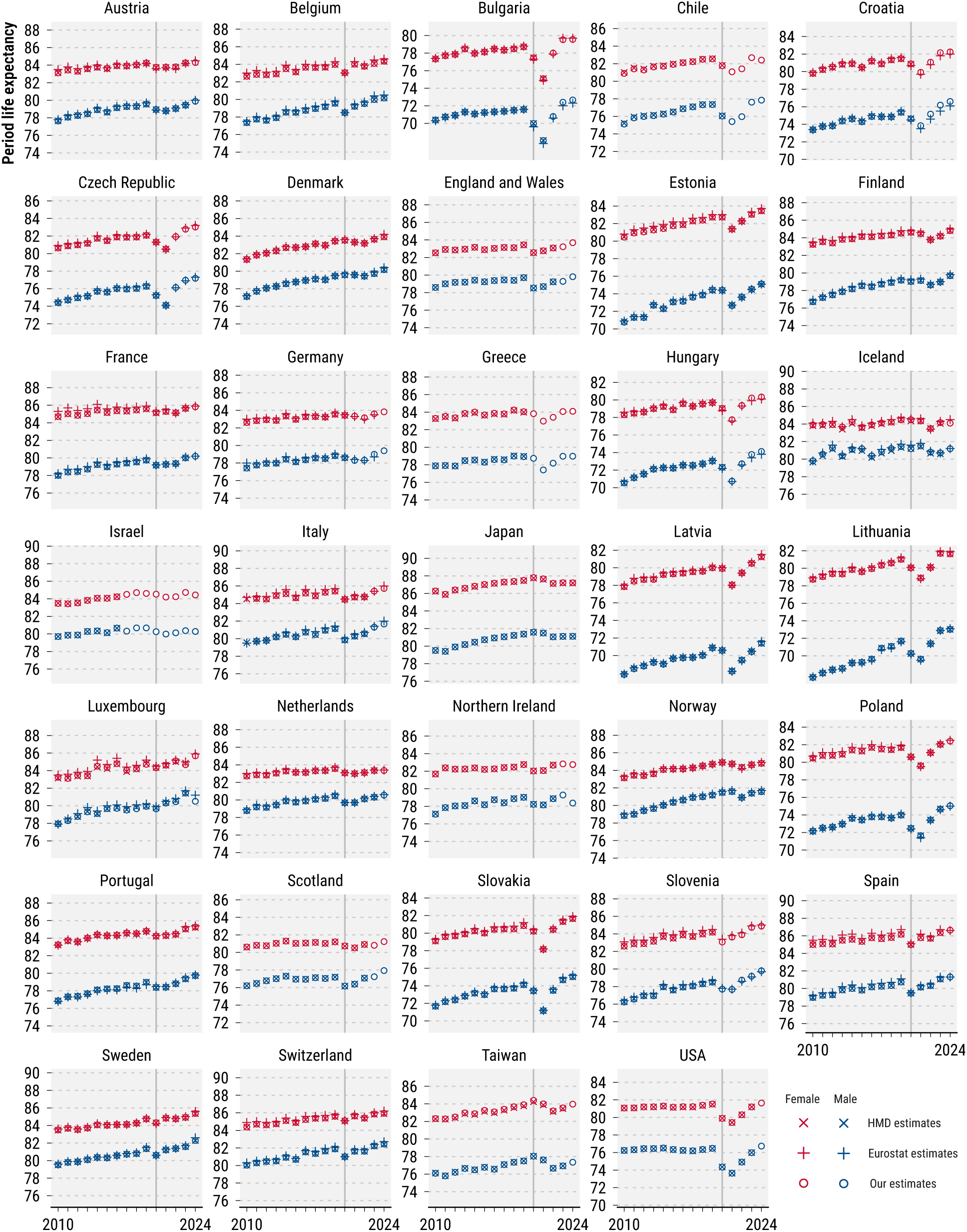
Comparison of our life expectancy estimates with past estimates by Eurostat and the Human Mortality Database.

### 4.2 Alternative measures of excess mortality

In addition to the life expectancy deficit we calculate^9^ alternative measures of excess mortality as a sensitivity check.

Excess deaths are the difference between actual and expected deaths,

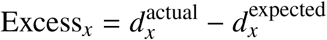

and can be turned into the P-score[11] by aggregating over age and expressing them as a ratio to total expected deaths,

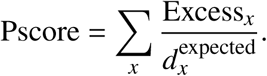

The mean unfulfilled lifespan (MUL, [12]), like the life expectancy deficit, is based on counterfactual life expectancies. MUL is defined as the difference between the average age at death during some period and the average expected age at death under counterfactual mortality conditions.

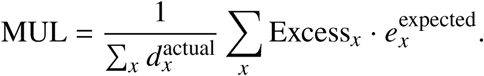

Figure 5 shows a comparison of the three measures with each other across countries.

**Figure 5:**
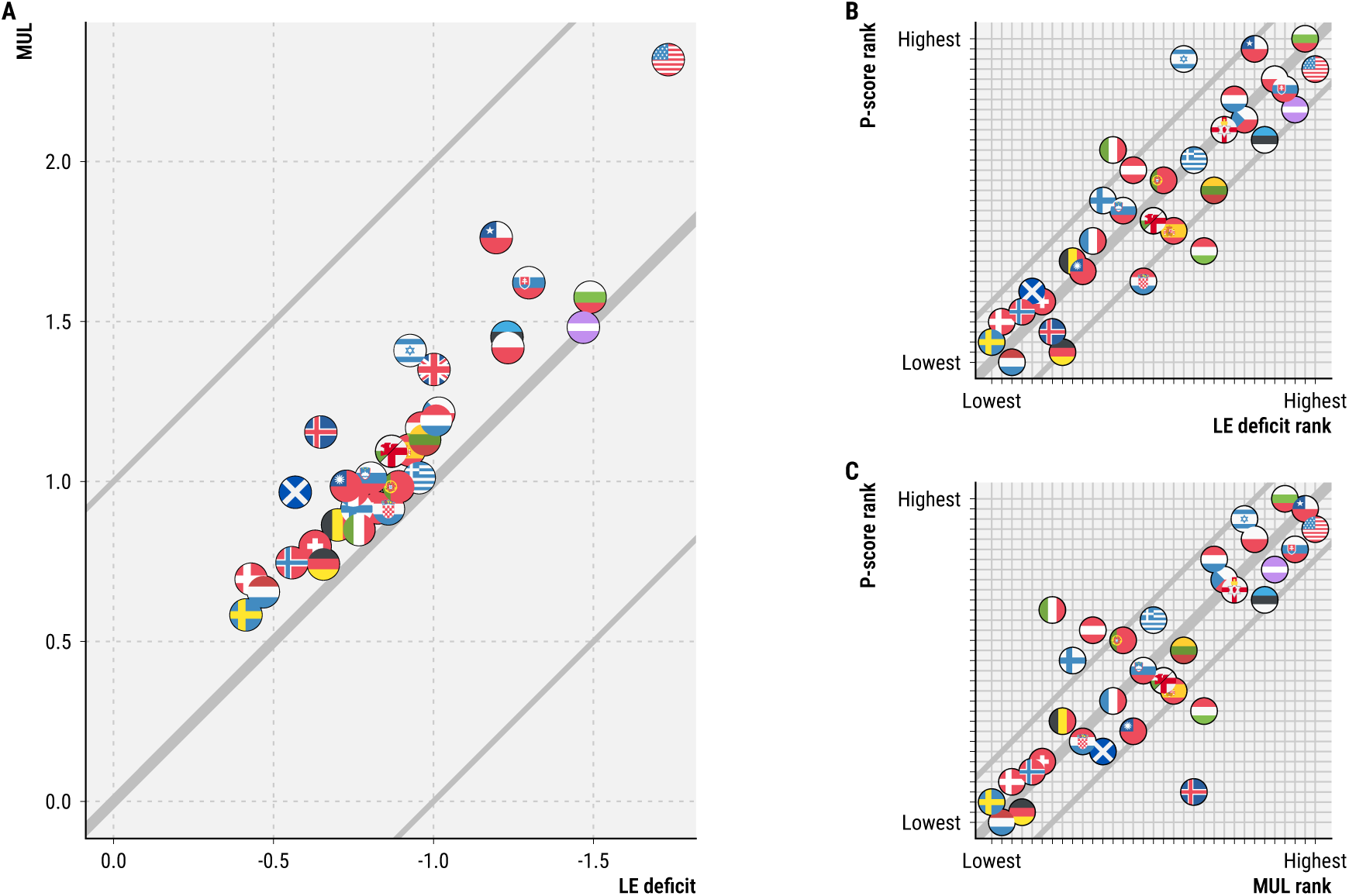
Cross-country comparison of alternative excess mortality measures for the period 2020-2024. A) Life expectancy deficit vs. mean unfulfilled lifespan, B) Country ranking of life expectancy deficit vs. P-score, C) Country ranking of mean unfulfilled lifespan vs. P-score.

### 4.3 Sensitivity of results to choice of fitting period

As a robustness check we recalculated key results in our paper under a 10-year fitting period for the counterfactual life expectancy. Key results remain robust under either 10 or 20 years of fitting while the existence of significant life expectancy deficits in 2024 changes for some countries.

The cluster assignment under the hierarchical clustering algorithm is perfectly robust against changes in the fitting period with identical clusters identified under either analysis. Visually, the salient features of each cluster remain robust. A 10-year fitting period yields slightly lower life expectancy deficits 2020-2024 compared to the longer fit (see Figures 6 and 7).

**Figure 6:**
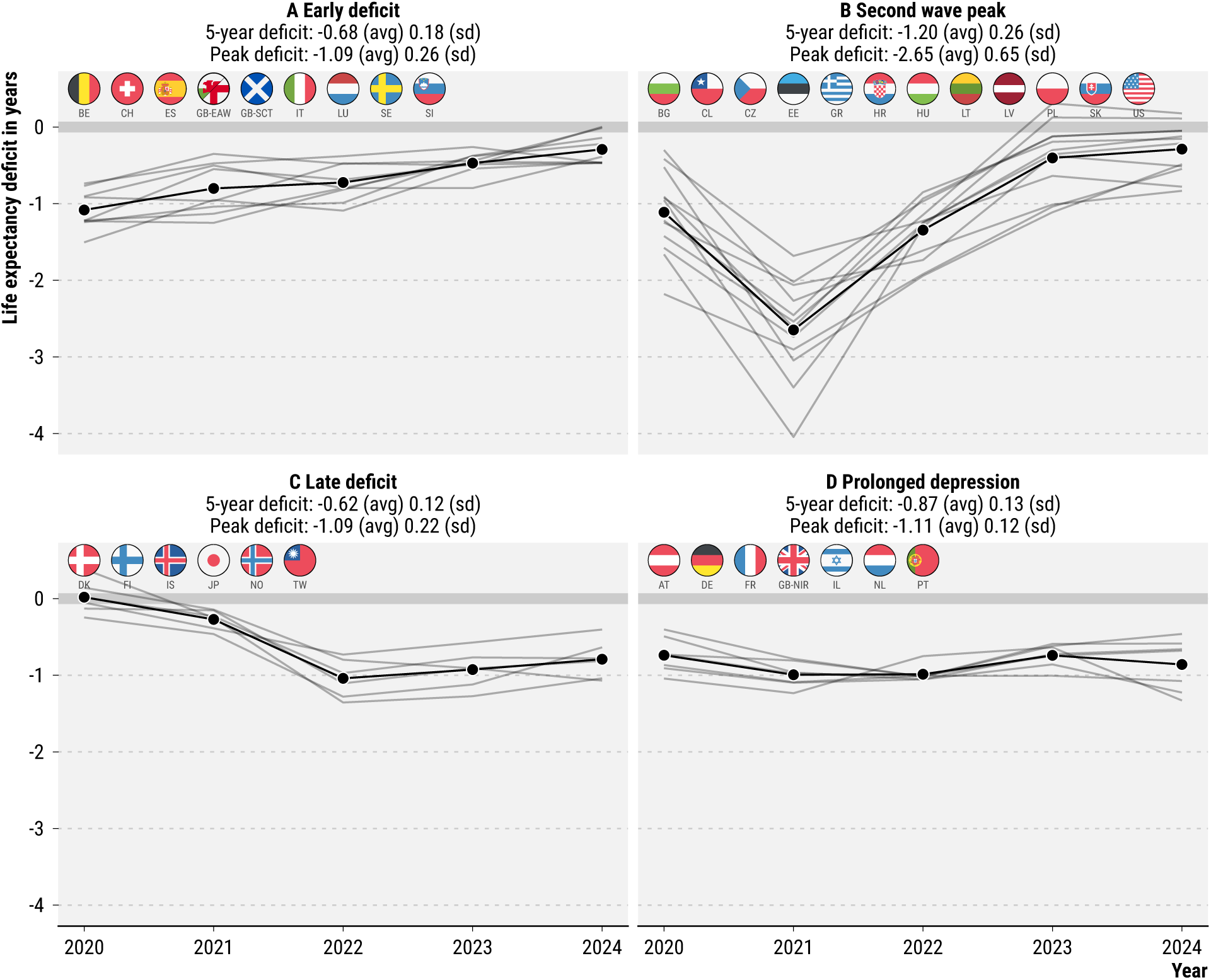
Deficit typology under 20-year Lee-Carter fitting period.

**Figure 7:**
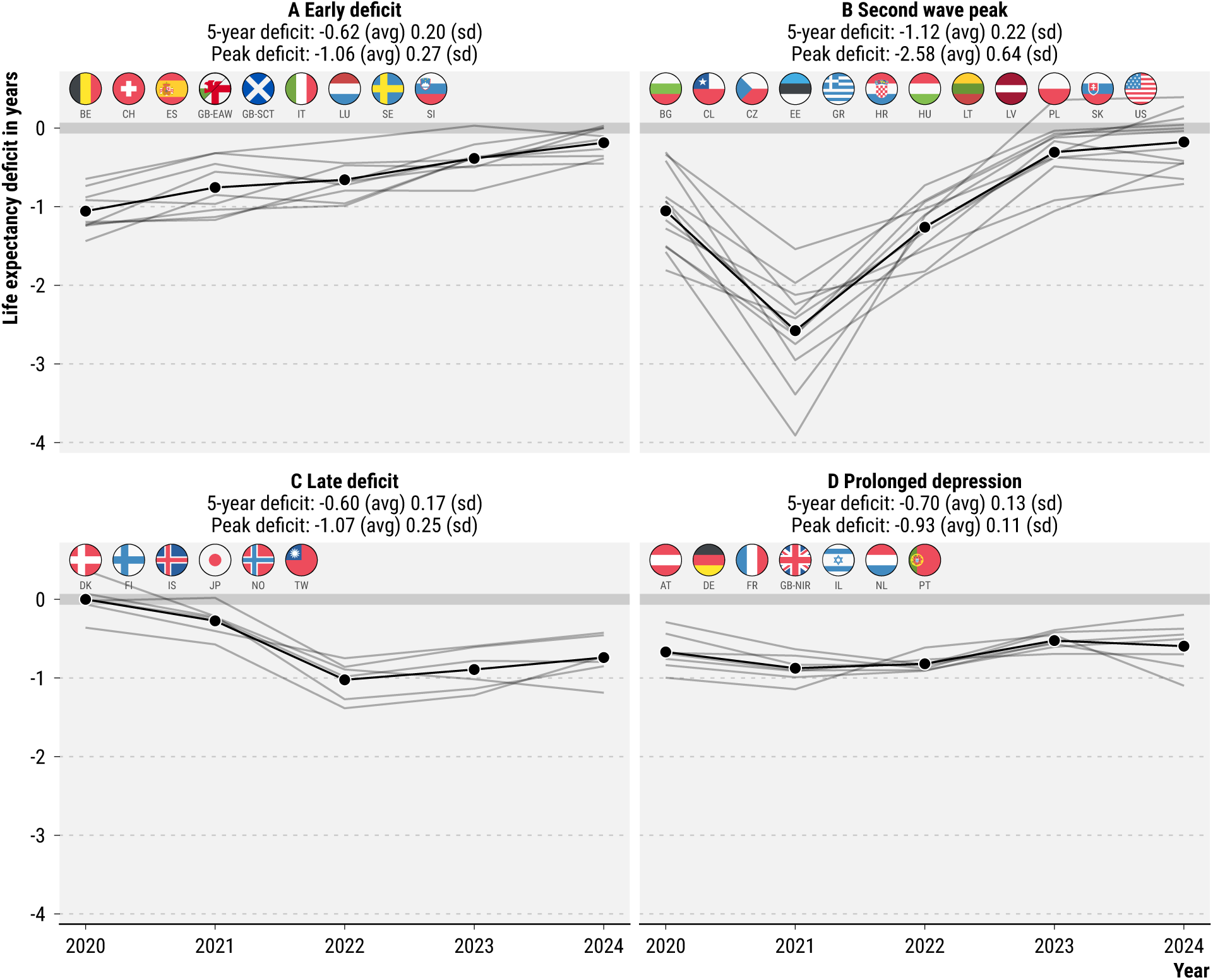
Deficit typology under 10-year Lee-Carter fitting period.

Under a 20-year fitting period we estimated 12 countries (AT, CL, DK, FI, GB-NIR, GR, IL, JP, NL, NO, TW, US) with a significant (*p* < 0.05) life expectancy deficit in 2024 compared to 9 (CL, DK, FI, GB-NIR, IL, JP, NL, NO, TW) under a 10-year fit.

The US is uniquely sensitive to the choice of fitting period due to a period of life expectancy stagnation pre-2020. A 10-year fitting window continues the flat trajectory, leading to lower life expectancy deficits, whereas a 20-year fitting window extrapolates a longer-term trend of slight life expectancy increase. As a result the finding that the US had the highest deficit overall is not robust (see Figures 8 and 9).

**Figure 8:**
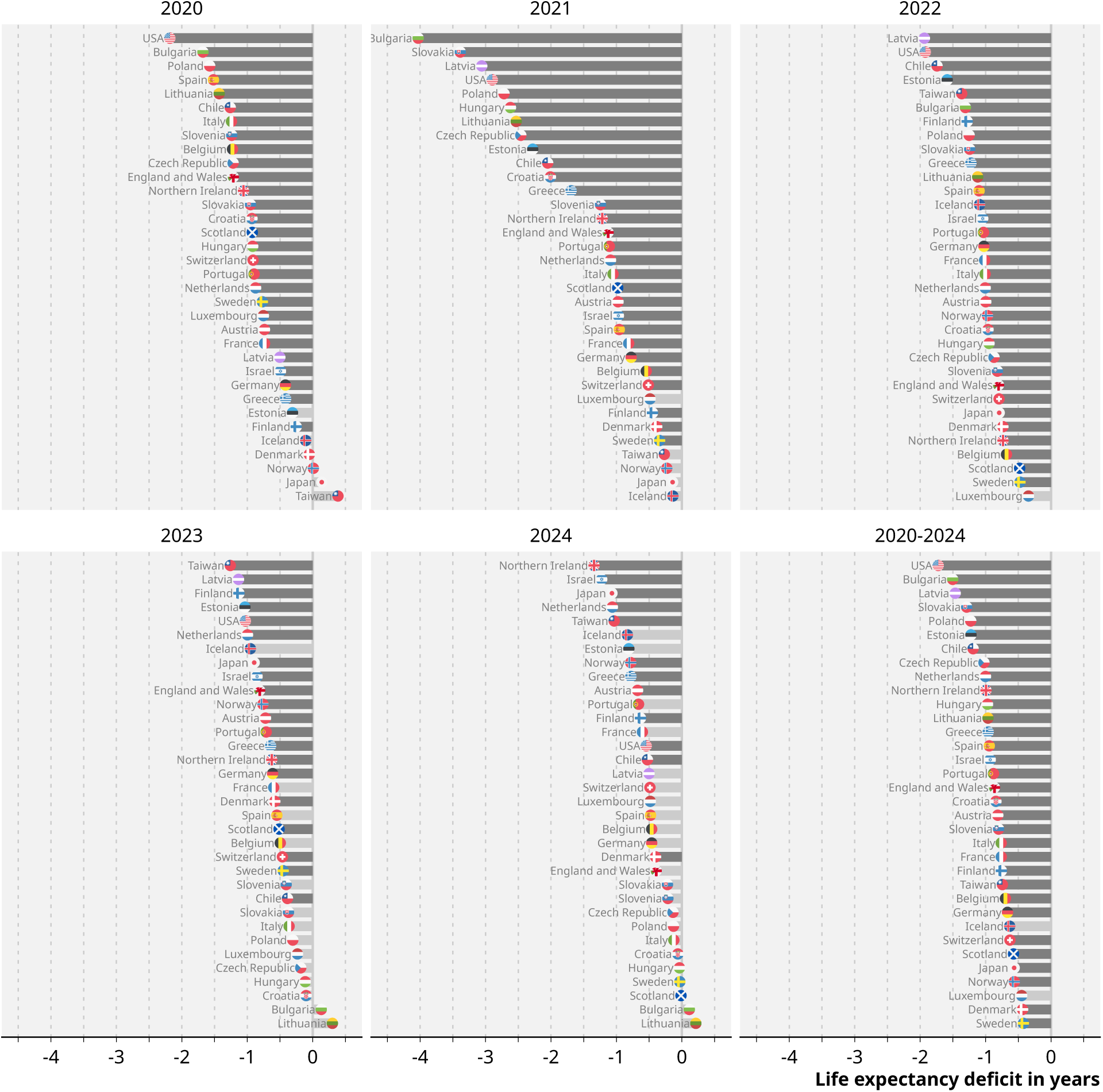
Annual life expectancy deficits under 20-year Lee-Carter fitting period.

**Figure 9:**
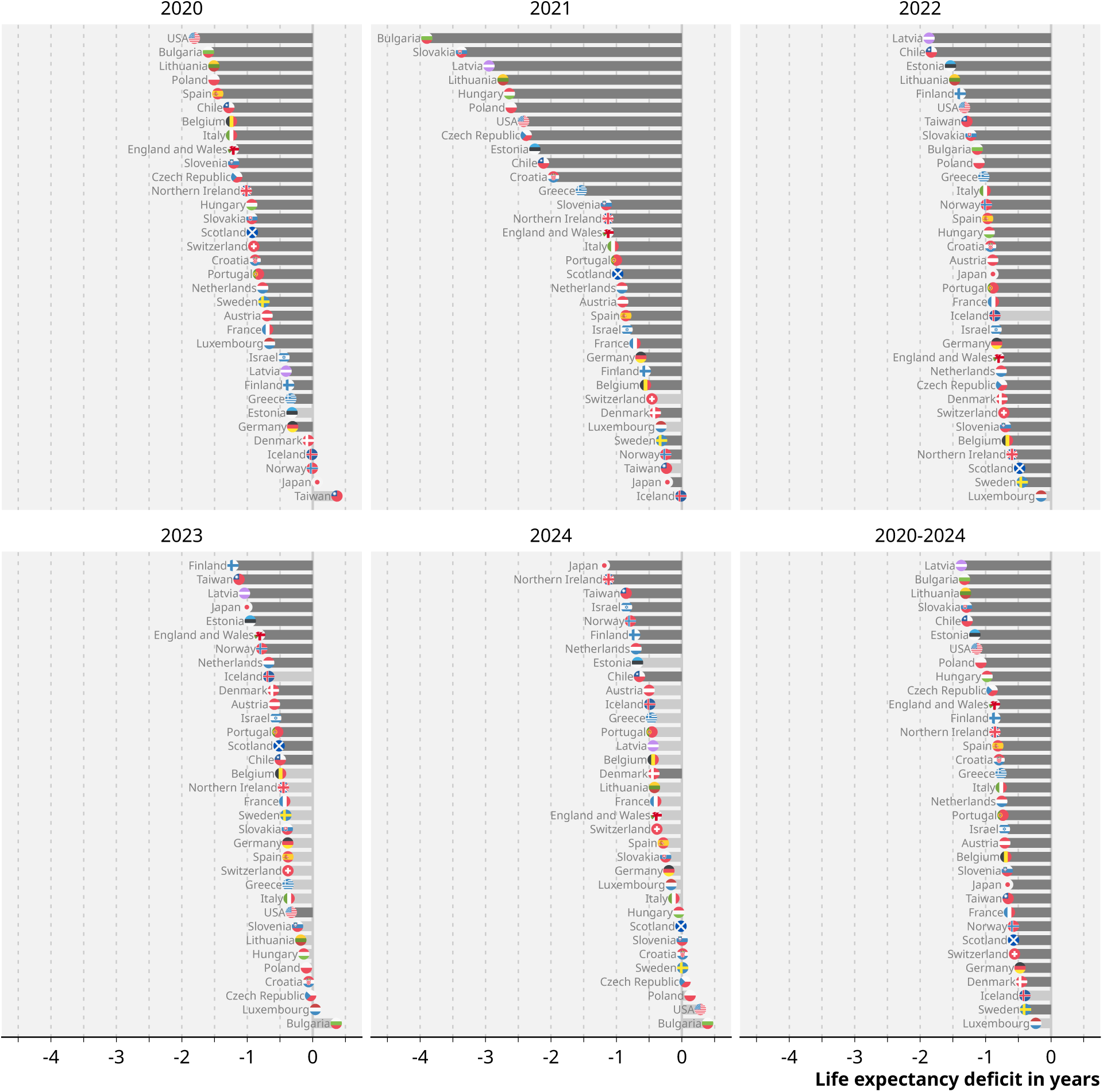
Annual life expectancy deficits under 10-year Lee-Carter fitting period.

### 4.4 Sensitivity of results to choice of counterfactual

The choice of model for producing the counterfactual life expectancy under continuation of prepandemic trends influences life expectancy deficit estimates. Here we compare^10^ our results under three different counterfactuals a) the Lee-Carter forecasts used in the main text, b) a direct extrapolation of 2019 life expectancy under continuation of the mean annual changes observed over 2000-2019 (LE random walk with drift), and c) a log-linear extrapolation of age-specific death rates based on mean annual changes 2000-2019 (ASDR random walk with drift).

Figure 10 displays the counterfactual life expectancy forecasts under different models. For the majority of locations the results are coherent. In countries where forecasts diverge noticeably, such as Estonia, Iceland, Israel, the Netherlands, Scotland, and the US, the Lee-Carter model is always the least optimistic one, i.e. it produced lower counterfactual life expectancies compared with the alternative approaches. This tendency of the Lee-Carter model to be conservative in its forecasts makes our findings of life expectancy deficits conservative as well. The deficits we find tend to be even larger under alternative models.

**Figure 10:**
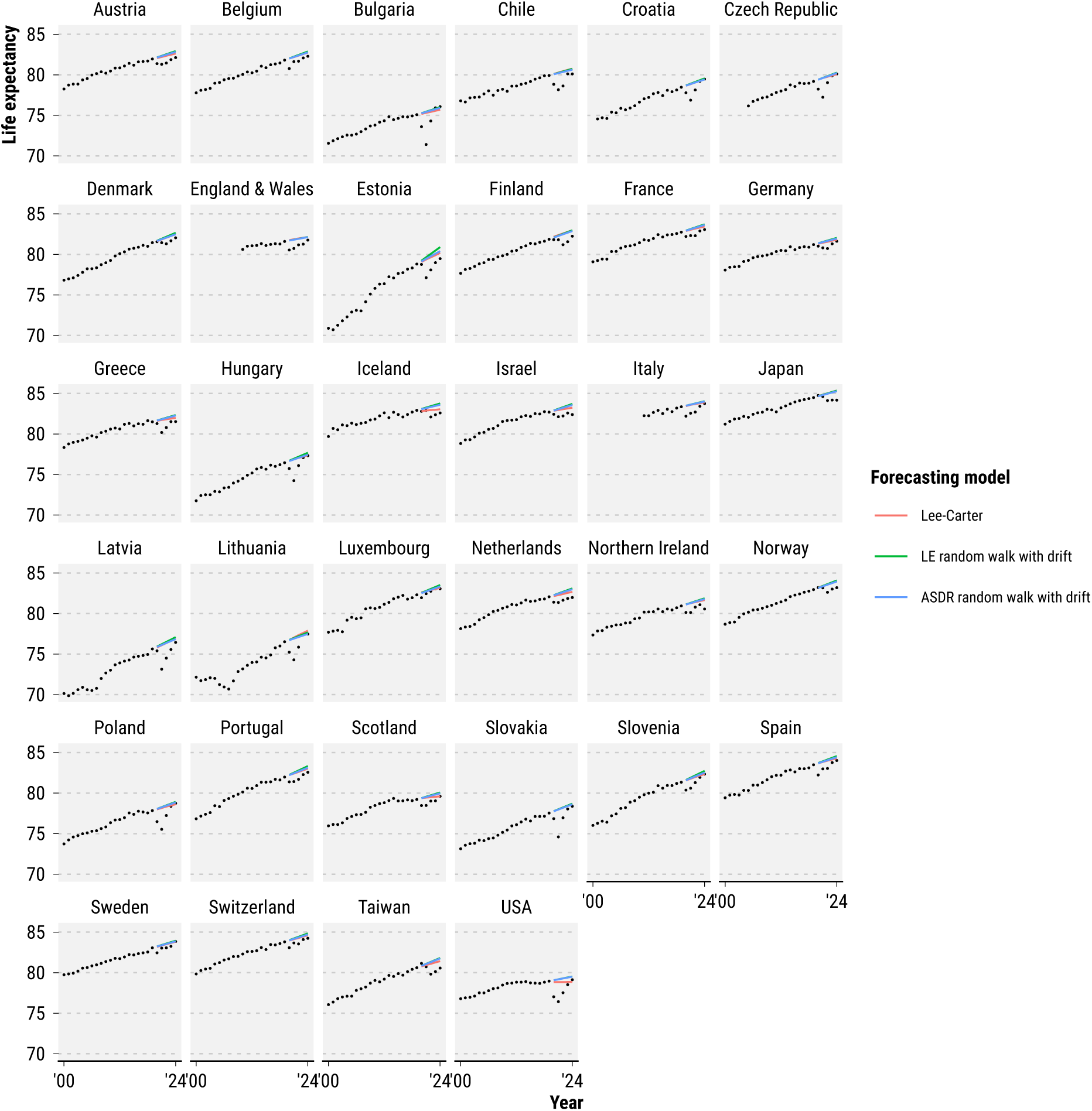
Comparison of life expectancy forecasts 2020-2024 under three different models.

The country ranking in annual life expectancy deficits is somewhat sensitive to the choice of counterfactual model and becomes more sensitive over time as shown in Figure 11. However, almost all countries are within ±5 ranks of their Lee-Carter rank when comparing against the two alternative counterfactuals. Notable exceptions are the US in 2023 and 2024 and Iceland in 2022. These changes in rank under different counterfactual life expectancy trajectories are, however, consistent with the prediction intervals given by the Lee-Carter method, which also derive from a set of simulated plausible counterfactuals. The prediction intervals around life expectancy deficits are wide enough to reliably identify the top, middle, and bottom groups in cross-country comparisons of life expectancy deficits, but do not allow for the robust identification of exact ranks.

**Figure 11:**
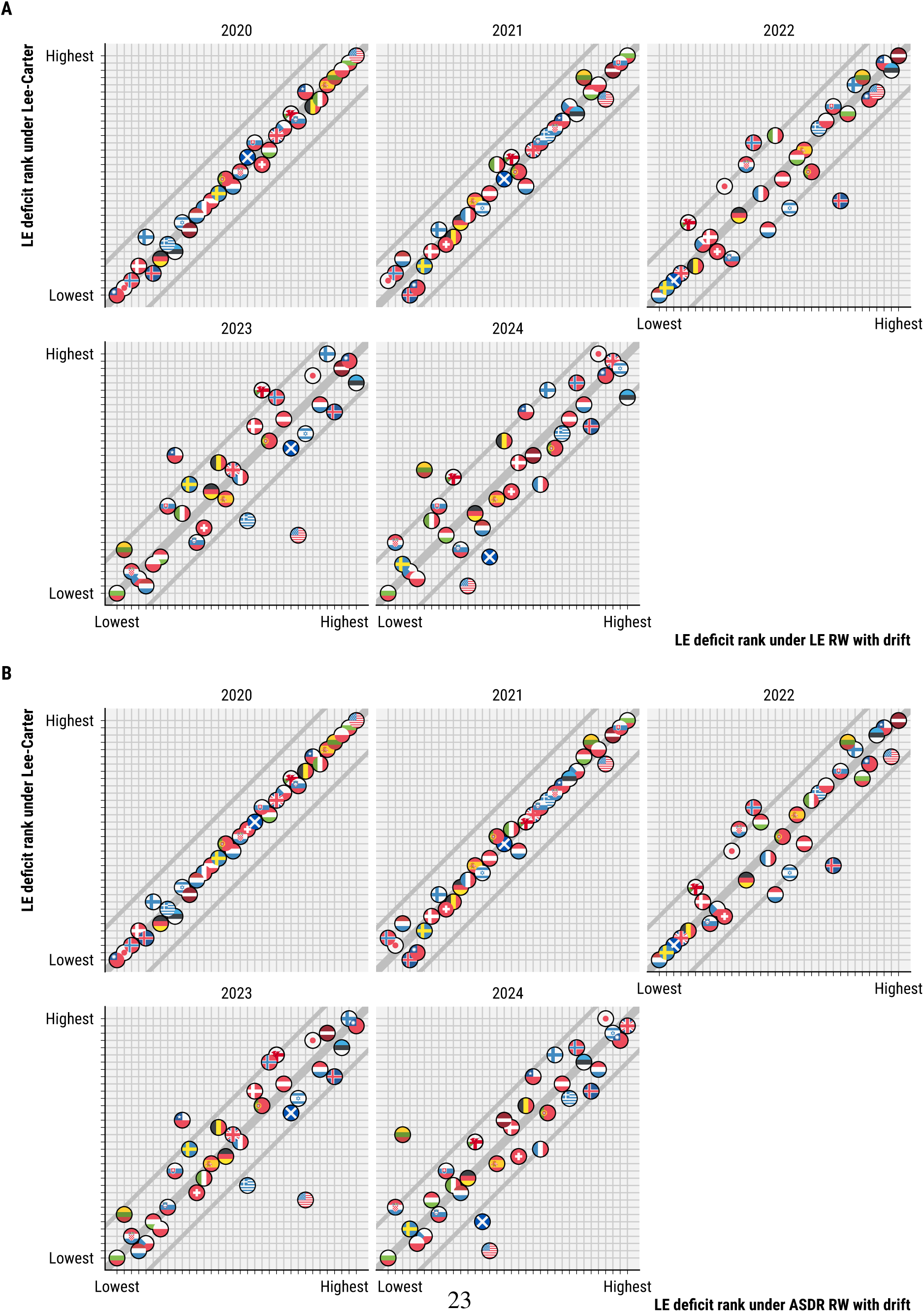
Country ranking of annual life expectancy deficits under different counterfactual models. Comparison of the Lee-Carter counterfactual with A) a direct extrapolation of life expectancy under continuation of average annual rate of increase over the time period 2000-2019, and B) a log-linear extrapolation of age-specific death rates based on average changes over the time period 2000-2019.

In Figure 12 we apply the same clustering as in the main text to life expectancy deficits under each counterfactual. Clearly the main cluster features reproduce almost perfectly across the different counterfactuals.

**Figure 12:**
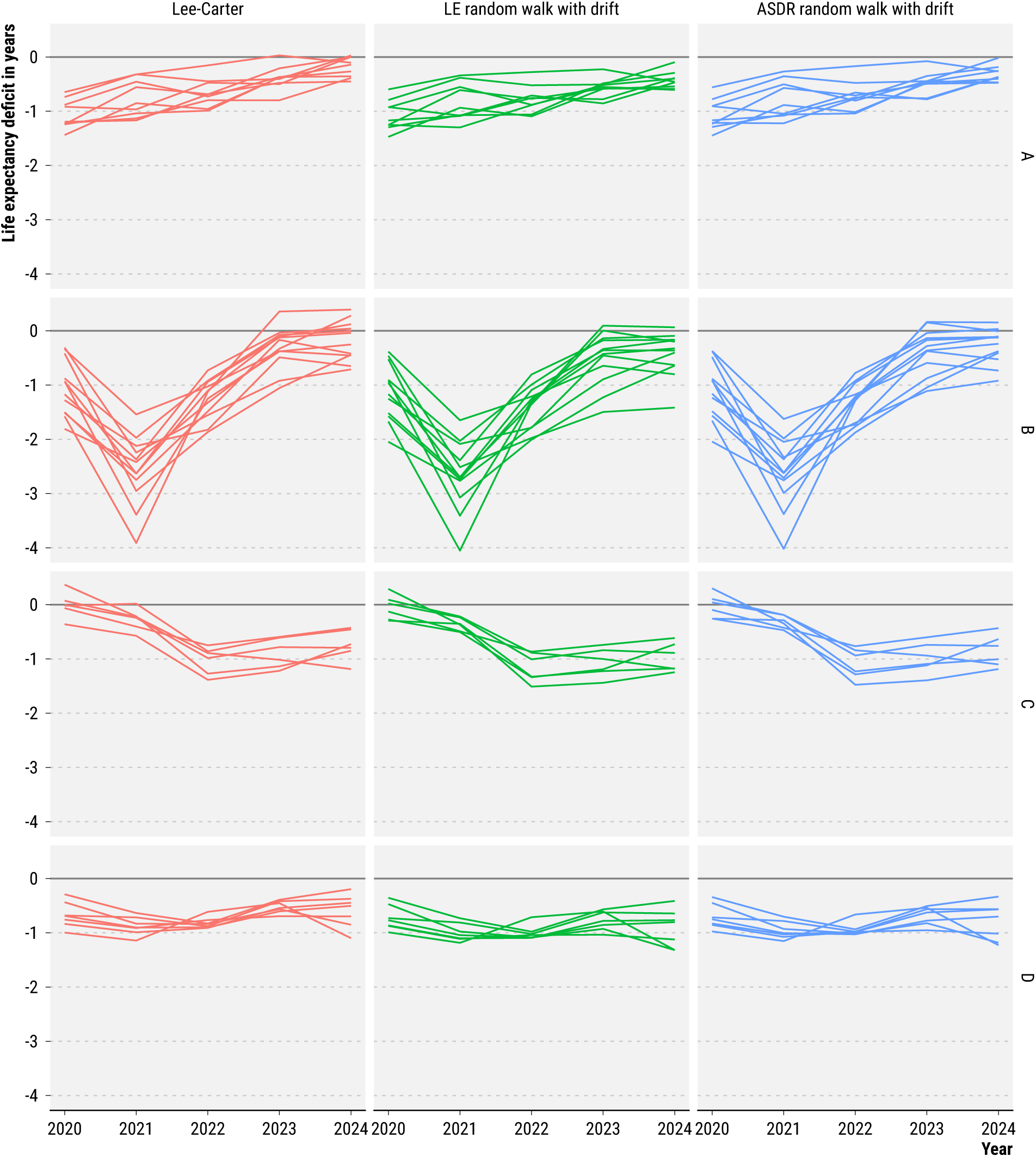
Annual life expectancy deficits and clusters under different counterfactual models.

### 4.5 Sensitivity of results to choice of clustering procedure

To establish robustness, we compare the group assignment under three different clustering procedures^11^.

#### Hierarchical clustering

The algorithm is the same as described in section 3.3 and as used in the main text.

#### Probabilistic decision rule

A similar cluster assignment as under the hierarchical algorithm can be achieved with a much simpler method of decision rules over our posterior simulation draws. First we test if a country’s life expectancy deficit series features a significant (*α* = 0.1) peak or slope. If no, we assign the group D, otherwise we check if the probability for 2021 being the peak deficit year is ≥ 90%. If yes, we assign the group B, otherwise we check if the mean peak deficit was later than 2021. If yes we assign group C, otherwise group A. This method achieves an overlap of 85.3% in cluster assignment with the hierarchical clustering method.

#### Non-probabilistic decision rule

An even simpler, entirely non-probabilistic, algorithm for cluster assignment can be defined by labeling as group D every country where the deficit peak is less than 0.2 years above the deficit in neighboring years and the range is less than 0.75 years. For all remaining countries the year of peak deficit determines group A (2020), group B (2021), or group C (later than 2021). This simplistic method assigns the same label as the hierarchical clustering method in 82.4% of regions.

Table 2 shows the cluster assignments per country under the three different clustering procedures. The results in the paper are reported under the hierarchical clustering.

**Table 2:** Regional cluster assignments under different clustering procedures.

| Region | Hierarchical | Probabilistic | Non-probabilistic |
| --- | --- | --- | --- |
| AT | D | D | D |
| BE | A | A | A |
| BG | B | B | B |
| CH | A | A | A |
| CL | B | B | B |
| CZ | B | B | B |
| DE | D | C | C |
| DK | C | C | C |
| EE | B | B | B |
| ES | A | A | A |
| FI | C | C | C |
| FR | D | C | C |
| GB-EAW | A | A | A |
| GB-NIR | D | C | C |
| GB-SCT | A | A | B |
| GR | B | B | B |
| HR | B | B | B |
| HU | B | B | B |
| IL | D | C | C |
| IS | C | C | C |
| IT | A | A | A |
| JP | C | C | C |
| LT | B | B | B |
| LU | A | D | A |
| LV | B | B | B |
| NL | D | D | D |
| NO | C | C | C |
| PL | B | B | B |
| PT | D | D | D |
| SE | A | A | A |
| SI | A | A | B |
| SK | B | B | B |
| TW | C | C | C |
| US | B | B | B |

## Footnotes

1 Per the documentation of the Human Mortality Database, “death and population data for Israel refer to all populations subject to Israeli law, which includes residents of East Jerusalem, the Golan Heights and Jewish settlements in the Palestinian territories occupied since 1967. The Human Mortality Database (HMD) takes no position concerning the current or future legal or political status of these areas.” We interpret this as meaning that deaths in Gaza and Palestinians in the West Bank are excluded.

1 github.com/jschoeley/e0deficit/blob/main/src/12-download_death. R github.com/jschoeley/e0deficit/blob/main/src/11-download_population_and_rates. R

2 https://github.com/jschoeley/e0deficit/blob/main/src/21-harmonize_death_with_exposures.R

3 https://github.com/jschoeley/e0deficit/blob/main/src/31-rate_bias_correction.R

4 https://github.com/jschoeley/e0deficit/blob/main/src/40-counterfactual_projection.R

5 https://github.com/jschoeley/e0deficit/blob/main/src/50-estimation_and_inference.R

6 https://github.com/jschoeley/e0deficit/blob/main/src/51-assign_clusters.R

7 https://github.com/jschoeley/e0deficit/blob/main/src/50-estimation_and_inference.R

8 https://github.com/jschoeley/e0deficit/blob/main/src/91-benchmark_check_e0.R

9 https://github.com/jschoeley/e0deficit/blob/main/src/92-sensitivity_check_excess_-measures.R

10 https://github.com/jschoeley/e0deficit/blob/main/src/95-sensitivity_check_model_choice.R

11 https://github.com/jschoeley/e0deficit/blob/main/src/51-assign_clusters.R

